# A cell type-aware framework for nominating non-coding variants in Mendelian regulatory disorders

**DOI:** 10.1101/2023.12.22.23300468

**Authors:** Arthur S. Lee, Lauren J. Ayers, Michael Kosicki, Wai-Man Chan, Lydia N. Fozo, Brandon M. Pratt, Thomas E. Collins, Boxun Zhao, Matthew F. Rose, Alba Sanchis-Juan, Jack M. Fu, Isaac Wong, Xuefang Zhao, Alan P. Tenney, Cassia Lee, Kristen M. Laricchia, Brenda J. Barry, Victoria R. Bradford, Monkol Lek, Daniel G. MacArthur, Eunjung Alice Lee, Michael E. Talkowski, Harrison Brand, Len A. Pennacchio, Elizabeth C. Engle

## Abstract

Unsolved Mendelian cases often lack obvious pathogenic coding variants, suggesting potential non-coding etiologies. Here, we present a single cell multi-omic framework integrating embryonic mouse chromatin accessibility, histone modification, and gene expression assays to discover cranial motor neuron (cMN) *cis*-regulatory elements and subsequently nominate candidate non-coding variants in the congenital cranial dysinnervation disorders (CCDDs), a set of Mendelian disorders altering cMN development. We generated single cell epigenomic profiles for ∼86,000 cMNs and related cell types, identifying ∼250,000 accessible regulatory elements with cognate gene predictions for ∼145,000 putative enhancers. Seventy-five percent of elements (44 of 59) validated in an *in vivo* transgenic reporter assay, demonstrating that single cell accessibility is a strong predictor of enhancer activity. Applying our cMN atlas to 899 whole genome sequences from 270 genetically unsolved CCDD pedigrees, we achieved significant reduction in our variant search space and nominated candidate variants predicted to regulate known CCDD disease genes *MAFB, PHOX2A, CHN1,* and *EBF3* – as well as new candidates in recurrently mutated enhancers through peak- and gene-centric allelic aggregation. This work provides novel non-coding variant discoveries of relevance to CCDDs and a generalizable framework for nominating non-coding variants of potentially high functional impact in other Mendelian disorders.

## INTRODUCTION

While the great majority of genetic variants associated with complex disease are common in the population and localize to non-coding sequences, less than 5% of the known Mendelian phenotype entries in OMIM have been attributed to non-coding mutations^1–4^. However, it remains unsettled the extent to which this disparity in coding:non-coding causal Mendelian variants is explained by the relative effect sizes of coding vs. non-coding variation, difficulty in deciphering the functional impact of non-coding variation, and/or ascertainment due to greater number and size of exome-versus genome-sequenced disease cohorts^1,5–8^. Nominating pathogenic non-coding variants in Mendelian disease remains a major challenge due to a vastly increased search space (98% of the genome) relative to coding variants. Compounding this challenge is the lack of a generalizable rubric for nominating non-coding pathogenic variants relative to the more readily interpretable molecular and biochemical constraints governing protein coding variant effects.

In recognition of these challenges, large-scale functional genomics projects such as ENCODE and Roadmap Epigenomics have provided valuable and expansive genome-wide functional information across a growing array of potentially disease-relevant tissues and cell types^9,10^. Such efforts reveal that the non-coding genome is abundant with *cis* regulatory elements (cREs) - segments of non-coding DNA that regulate gene expression through transcription factor binding and three-dimensional physical interactions with their cognate genes. Biologically active cREs are associated with accessible chromatin, and combinations of accessible cREs vary dramatically among different cell types^11^. Therefore, understanding the chromatin accessibility landscape of cell types affected in disease is critical to identifying and interpreting disease-causing variation in the non-coding genome.

Disease-relevant developmental processes are disproportionately driven by regulation of gene expression^12,13^, making congenital genetic disorders attractive candidates for non-coding etiologies. However, sampling developing human cell types remains particularly challenging, as samples are often restricted by cell location, assayable cells, invasiveness of sampling, and/or extremely narrow windows of biologically-relevant regulation of gene expression and development^14^. Thus, while fetal epigenomic reference sets are emerging for humans, samples are generally assayed at the whole-organ/tissue level and/or at later stages of development, making appropriate sampling and identification of early-born and rare cell types difficult^15^. By contrast, sample collection and marker-based enrichment in model organisms can achieve substantial representation of disease-relevant cell types at early stages of development^16–18^.

The congenital cranial dysinnervation disorders (CCDDs) are Mendelian disorders in which movement of extraocular and/or cranial musculature are limited secondary to errors in the development of cranial motor neurons (cMNs) or the growth and guidance of their axons (**Figure 1a**). Although a known subset of the CCDDs are caused by Mendelian protein-coding variants^19–28^, a substantial proportion of cases remain unsolved by whole exome sequencing, including pedigrees with Mendelian inheritance patterns and cases with classic phenotypic presentations lacking corresponding mutations in the expected genes (representing potential locus heterogeneity)^29^. Moreover, most CCDD cases are sporadic or segregate in small dominant families for which non-coding variant prioritization is extremely difficult.

**Figure 1.**
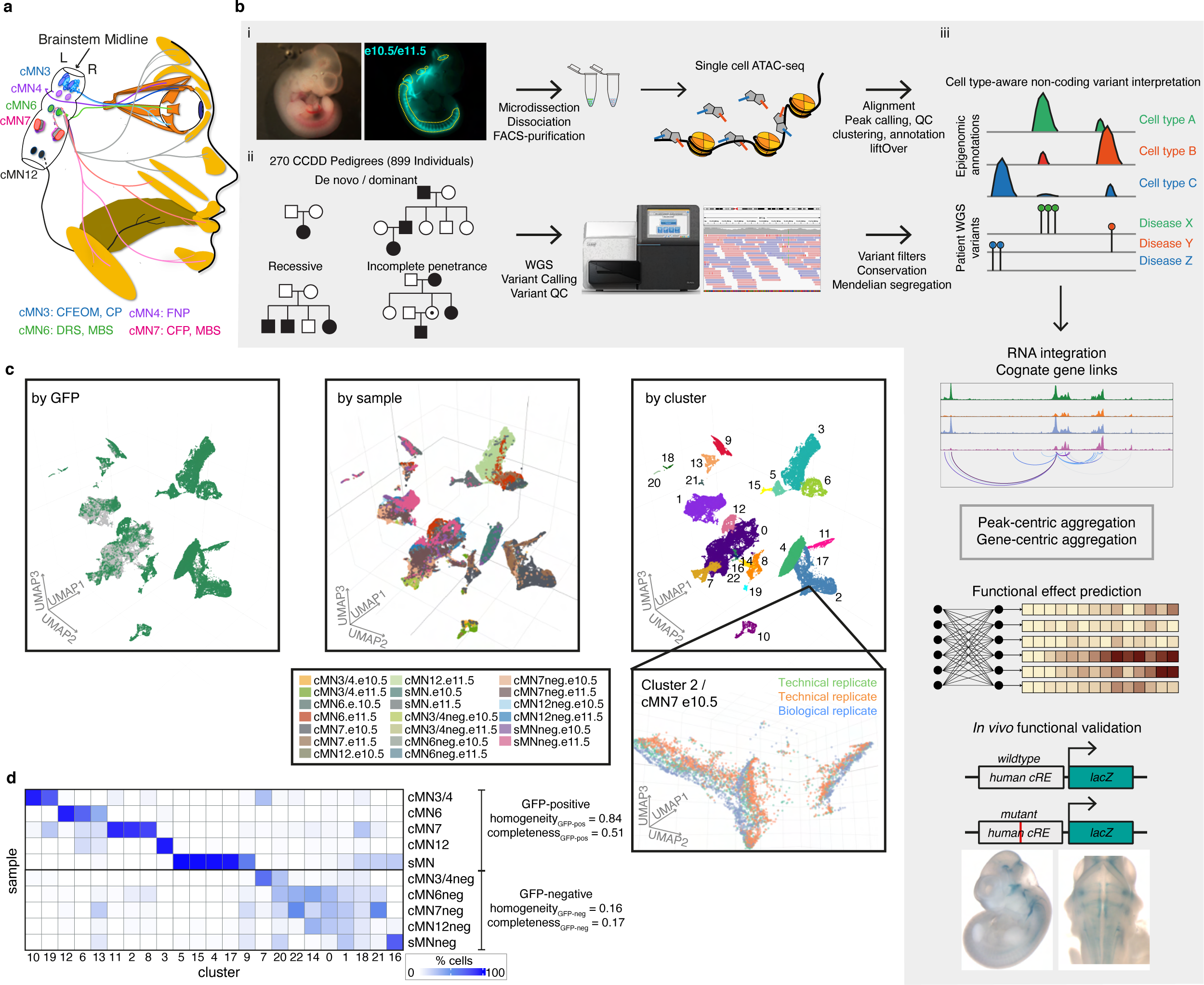
Integrating Mendelian pedigrees with single cell epigenomic data. a. Schematic depicting subset of human cMNs and their targeted muscles. cMN3 (blue) = oculomotor nucleus which innervates the inferior rectus, medial rectus, superior rectus, inferior oblique, and levator palpebrae superior muscles; cMN4 (purple) = trochlear nucleus which innervates the superior oblique muscle; cMN6 (green) = abducens nucleus which innervates the lateral rectus muscle (bisected); cMN7 (pink) = facial nucleus which innervate muscles of facial expression; cMN12 (black) = hypoglossal nucleus which innervates tongue muscles. Corresponding CCDDs for each cMN are listed under diagram and color coded. CFEOM: congenital fibrosis of the extraocular muscles; CP: congenital ptosis; FNP: fourth nerve palsy; DRS: Duane retraction syndrome; MBS: Moebius syndrome; CFP: congenital facial palsy. b. Overview of the experimental and computational approach. i) Generating cell type-specific chromatin accessibility profiles. Brightfield and fluorescent images of e10.5 *Isl1^MN^*:GFP embryo (left) from which cMNs are microdissected (yellow dotted lines, dissociated, FACS-purified (middle), followed by scATAC and data processing (right; red and blue lines represent adapters, black line represents DNA, orange cylinders represent nucleosomes, grey pentagons represent Tn5). ii) WGS of 270 CCDD pedigrees (left; 899 individuals; sporadic and inherited cases) followed by joint variant calling, QC, and Mendelian variant filtering (right). iii) Integrating genome-wide non-coding variant calls with epigenomic annotations for variant nomination (top). To aid in variant interpretation, we identify cognate genes (2^nd^ row), aggregate candidate variants, generate functional variant effect predictions (3^rd^ row), and validate top predictions *in vivo* (bottom). c. UMAP embedding of single cell chromatin accessibility profiles from 86,089 GFP-positive cMNs, sMNs, and their surrounding GFP-negative neuronal tissue colored based on GFP reporter status (left, GFP-positive green, GFP-negative grey), sample (middle, with sample key under UMAP) and cluster (right, with cluster annotations in **Supplementary Table 3**). Gridlines in middle UMAP apply to left and right UMAPs as well. The inset shows the relative proximity of Cluster 2 cells dissected from the same cell type (cMN7 e10.5) from different technical and biological replicates. d. Heatmap depicting the proportions of dissected cells within each of the 23 major clusters. Homogeneity/completeness metrics are shown for GFP-positive versus GFP-negative clusters. cMN6 and cMN7 are in close spatial proximity and are commonly co-dissected.

The CCDDs represent an attractive test case for dissecting cell type-specific disorders, as defects in specific cMN populations are highly stereotyped with predictable corresponding human phenotypes^30^. By contrast, many complex and even some Mendelian diseases are not immediately attributable to an unambiguous, singular cell type of interest, making assaying appropriate cell types a major challenge^31–33^. Moreover, while sampling and identification of developing cMNs at disease-relevant timepoints is extremely difficult in developing human embryos, cMN birth, migration, axon growth/guidance, and mature anatomy/nerve branches are exquisitely conserved between humans and mice^30^. Motor neuron reporter mice permit sample collection and marker-based enrichment of cMNs at these key stages of development. Importantly, we have previously demonstrated that such mouse models helped to characterize non-coding pathogenic variants that alter gene expression in HCFP1, a disorder of facial nerve (cMN7) development^34^. Here, to comprehensively discover the repertoire of cREs underlying proper cMN development, we have generated a chromatin accessibility atlas of developing mouse cMNs and adjacent cell types. We subsequently use this atlas to reduce our candidate variant search space and ultimately interpret and nominate non-coding variants among 270 unsolved CCDD pedigrees (**Figure 1b, Supplementary Table 1**).

## RESULTS

### Defining disease-relevant cREs in the developing cMNs

To discover disease-relevant cREs and ultimately reduce our non-coding search space for nominating candidate pathogenic CCDD variants, we generated a single cell atlas of embryonic mouse cMN chromatin accessibility. Using wildtype or transgenic mice expressing GFP under the *Isl1^MN^*:GFP or *Hb9*:GFP motor neuron reporters^35,36^ (**Figure 1ai**), we performed fluorescence-assisted microdissection and FACS-based enrichment of GFP-positive primary mouse embryonic oculomotor (cMN3), trochlear (cMN4), abducens (cMN6), facial (cMN7), hypoglossal (cMN12), spinal motor neurons (sMNs), and surrounding GFP-negative non-motor neuron cells (-”neg”), followed by droplet-based single cell ATAC-seq (scATAC). cMN birth and development occur continuously over a period of weeks in early human embryos and days (e9.0-e12.5) in mice^34,37^. For the known CCDD genes, mRNA expression and/or observed cellular defects typically overlap key developmental timepoints e10.5 and e11.5 in mice – both for cellular identity-related transcription factor^38–42^ and axon guidance-related^22,43,44^ variants. Therefore, we captured these two embryonic timepoints for each cMN sample, reasoning that a major proportion of relevant cellular birth and initial axonal wiring would be represented at these ages^34,37^. At these stages, these cranial nuclei contain only hundreds (cMN3, 4, 6) to thousands (cMN7, 12) of motor neurons per nucleus, per embryo^43–45^.

We generated scATAC data across 20 unique sample types (cMN3/4, 6, 7, 12, and sMN for GFP-positive and -negative cells, each at e10.5 and e11.5), 9 with biological replicates and 2 with technical replicates for 32 samples in total and sequenced them to high coverage (mean coverage = 48,772 reads per cell). We included GFP-negative cells to reduce uncertainty in peak calling, further increase representation from rare cell types, and capture regional-specific cell types that could harbor elements conferring non-cell-autonomous effects on cMN development. To generate a high-quality set of non-coding elements, we performed stringent quality control (**Extended Data Figure 1a-h, Methods**). Altogether, we generated high-quality single-cell accessibility profiles for 86,089 (49,708 GFP-positive and 36,381 GFP-negative) cells, in some cases achieving substantial oversampling of cranial motor neurons in the developing mouse embryo (up to 23-fold cellular coverage). Our final dataset revealed prominent signals of expected nucleosome banding, a high fracton of reads in peaks (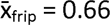), transcription start site enrichment, and strong concordance between biological replicates (**Figure 1c, Extended Data Figure 1d-h, Supplementary Table 2**). In addition to evaluating per-sample and per-cell metrics, we estimated a decrease in global accessibility over developmental time, consistent with observations in other developing cell types (*β_time_* = 0.049, p-value < 1 x 10^-15^, linear regression, **Supplementary Note 1**)^46,47^.

We performed bulk ATAC on a subset of microdissected and FACS-purified cMN samples to evaluate the concordance between bulk and single cell peak representation. As expected, bulk and single cell cMN ATAC peaks are highly correlated in their matching dissected cell types (**Extended Data Figure 2a,b**). scATAC peaks were enriched for intronic/distal annotations (relative to exonic/promoter annotations, OR = 1.9, p-value < 2.2 x 10^-16^, Fisher’s exact test) compared to bulk ATAC intronic/distal annotations, thus better capturing regions that harbor the overwhelming majority of regulatory elements (**Extended Data Figure 2c**)^48^. Next, to test the cellular resolution of our scATAC data, we leveraged differences in the strategies used for bulk (cMN3 without cMN4) vs. scATAC dissection (cMN3 and cMN4 combined) and performed cluster analysis on cMN3/4 samples only (*ad hoc* clusters C1-C20, **Extended Data Figure 2a,d,e**). We identified significant overlap between *ad hoc* clusters C18 and C20 scATAC peaks with bulk cMN3 peaks. Moreover, we confirmed accessibility of known cMN3 markers in C18 and C20, and cMN4 markers in C19^49,50^ (**Extended Data Figure 2e**). When comparing the scATAC peaks to bulk ATAC peaks in ENCODE^9^ sampled from major developing brain regions (forebrain, midbrain, hindbrain) at comparable timepoints, we observed diminished overlap for GFP-positive cMN samples relative to GFP-negative samples (**Extended Data Figure 3a**). Further stratifying scATAC peaks based on cell type specificity scores^51^ revealed that highly specific scATAC peaks had consistently lower bulk coverage than peaks with low specificity (**Extended Data Figure 3b,c**), consistent with findings that cell-type specific regulatory elements often act within small populations of cells and may be more difficult to capture and annotate with bulk methods^52,53^.

To further distinguish between rare, distinct cell types, we adopted an iterative clustering strategy (**Methods**)^51^. We first identified 23 major clusters that correspond with “ground truth” dissected cell types based on known anatomy (**Figure 1c,d**; **Supplementary Table 3**). Overall, GFP-positive clusters demonstrated much more uniform sample membership than GFP-negative clusters, as reflected by their differences in cluster homogeneity^54^ (*h_gfp-positive_* = 0.84 vs. *h_gfp-negative_* = 0.16) and purity metrics (**Figure 1d, Extended Data Figure 4a, Supplementary Table 4, Methods**). Upon examining differentially accessible genes and elements through manual curation, review of the literature, and gene ontology analysis, we assigned provisional cell identities to the 23 major clusters, of which 10 clusters are cranial and 5 are spinal motor neurons based on dissection origin, and 9 are cranial and 4 are spinal motor neurons based on putative annotation (**Supplementary Table 3**). To further resolve the heterogeneity within clusters and to identify functionally and anatomically coherent subpopulations, we performed iterative clustering^51^ on each major cluster and identified 132 unique subclusters (**Extended Data Figure 4bi,ii**). Of these, 59 have GFP-positive membership > 90%, representing highly pure motor neuron populations (**Extended Data Figure 4c**). We observe even more distinct anatomic/temporal membership at the subcluster level, particularly for GFP-negative samples (subcluster homogeneity *h_gfp-positive_* = 0.87 vs. *h_gfp-negative_* = 0.43). These findings are consistent with highly dynamic and proliferative neurodevelopmental processes during this time period^12^. Neither major cluster nor subcluster membership was driven by experimental batch (**Extended Data Figure 4d**).

### cMN cRE functional conservation between mouse and human

Common disease risk loci tend to overlap non-coding accessible chromatin in their corresponding cell types - including accessible chromatin that is more readily ascertained in mouse versus human tissues^15,51^. However, with the exception of a few exemplary elements (e.g., see refs ^55–57^), the extent of overlap between human/mouse elements underlying Mendelian traits is largely unknown. Therefore, to evaluate the functional conservation of cREs in our cranial motor neuron atlas, we performed *in vivo* humanized enhancer assays on a curated subset (n = 26) of our candidate scATAC peaks that were absent from the VISTA enhancer database^58^ and had peak accessibility/specificity in cMNs and general signatures of enhancer function (i.e., evolutionary conservation and non cMN-specific histone modification data^59^, **Supplementary Table 5, Methods**). These results validated our approach, as we detected positive enhancer activity (any reporter expression) in 65% (17/26) of candidates. Moreover, 11 of the 17 validated enhancers (65%, 42% overall) recapitulate the anatomic expression patterns (motor neuron expression) predicted from the scATAC accessibility profiles to the resolution of individual nuclei/nerves. By contrast, of 3,229 total non-coding elements assayed in the VISTA enhancer database, only 67 (2.1%) show reproducible evidence of enhancer activity in the cMNs. Thus, high quality single cell accessibility profiles are highly predictive of cell type specific regulatory activity.

### Motif enrichment and footprinting reveal putative cMN regulators

To identify transcription factors/motifs responsible for cell type identity, we performed motif enrichment and aggregated footprinting analysis across all 23 major clusters and identified both known lineage-specific motif enrichment as well as new potential cMN transcription factor/motif relationships (**Figure 2a,b**). For example, we identified significant motif and footprinting enrichment of midbrain transcription factor OTX1 in populations corresponding to developing oculomotor/trochlear motor neurons (cluster cMN3/4.10) and the midbrain-hindbrain boundary (cluster MHB.7)^60^. We also identified notable footprints for ONECUT2 in multiple motor neuron populations, including cMN3/4, cMN7, and putative pre-enteric neural crest-derived cells (clusters cMN3/4.19, cMN7.11, enteric.17; **Figure 2b**). Importantly, we detected positive footprint signals for known lineage-specific regulators such as JunD footprints in the spinal and lymphoid lineages^61,62^ (clusters sMN.15, WBC.18) and GATA1 footprints in the erythroid lineage^63^ (cluster RBC.20; **Figure 2b**). Due to the relatively high homogeneity across the motor neuron clusters, we also compared motif enrichment across broader anatomic/functional classes of motor neurons and brain regions (**Figure 2c**). We identified strong enrichment of regional markers such as DMBX1^64^ in midbrain samples (i.e., cMN3/4, cMN3/4neg). We also found motifs enriched among the ocular motor neurons (i.e., cMN3/4, cMN6) such as PAX5, providing new potential avenues for comparative studies.

**Figure 2.**
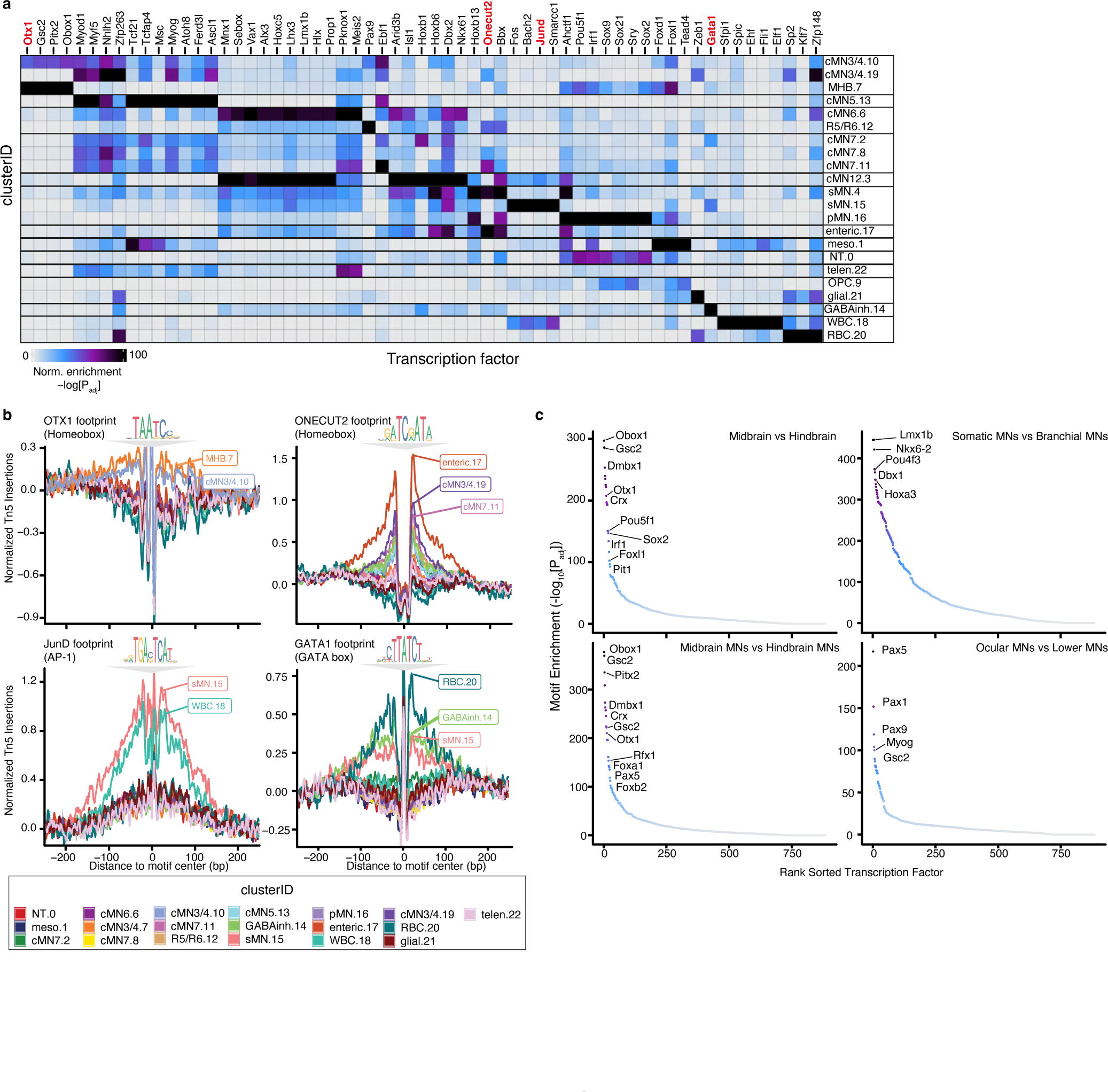
Motif enrichment and aggregate footprint analyses distinguish cell type specific TF binding motifs. a. Heatmap depicting enriched transcription factor binding motifs within differentially accessible peaks by cluster. Each entry is defined by its cluster identity (“clusterID.clusterNumber”). Corresponding cluster IDs and annotations are depicted. Color scale represents hypergeometric test p-values for each cluster and motif. Specific motifs and motif families vary significantly amongst clusters. Cluster annotations are defined in **Supplementary Table 3**. b. Aggregated subtraction-normalized footprinting profiles for a subset of cluster-enriched transcription factors (OTX1, ONECUT2, JunD, and GATA1) from (a), centered on their respective binding motifs. Specific clusters display positive evidence for TF motif binding for each motif. Corresponding motif position weight matrices from the CIS-BP database are depicted above each profile. Cluster IDs with corresponding color are below. c. Motif enrichment comparing broad classes of neuronal subtypes. Midbrain subtype contains motifs from cMN3/4neg cells; hindbrain from cMN6neg, cMN7neg, and cMN12neg cells; somatic MN from cMN3/4, cMN6, and cMN12 GFP-positive cells; branchial MN are from cMN7 GFP-positive cells; midbrain MN are cMN3/4 GFP-positive cells; hindbrain MN are cMN6, cMN7, and cMN12 GFP-positive cells; ocular MN are cMN3/4 and cMN6 GFP-positive cells; lower MN are cMN7, cMN12, and sMN GFP-positive cells. For each graph, the first listed subtype is enriched relative to the second listed subtype.

### Assigning cell type specific cREs to their cognate genes

A chief barrier to interpreting non-coding regulatory elements is identifying their *cis* target genes. While enhancers often regulate adjacent genes, many important regulatory links also occur over much longer distances, including known disease causing events^55,57,65–69^. Therefore, we generated scRNA data from GFP-positive and -negative cMN3/4, 6, and 7 at e10.5 and e11.5 (**Methods**) using reporter constructs, microdissection, and collection strategies analogous to those use used to generate the scATAC datasets. We then integrated these scRNA data with the cMN chromatin accessibility data to generate peak-to-gene links at the single cell level for putative cREs within +/- 500kb of a given gene (see **Methods**^70–72^). In total, we identified 145,073 known and putative enhancers with peak-to-gene links across the 23 clusters (median = 2 genes per enhancer, range = 1-37; **Supplementary Table 6**).

Because the accuracy of peak-to-gene links inferred from separate assays of ATAC and RNA data (“diagonal integration”)^73^ depends heavily on cell pairings, we performed multiple analyses to ensure that both our ATAC-RNA pairings and gene expression estimates were well calibrated. We compared our imputed single cell gene expression estimates to independently collected in-house bulk RNAseq experiments from cMN3, 4, 6, and 7 at e10.5 and e11.5 annotated with ground truth dissection labels (**Methods**). We identified strong positive concordance between imputed gene expression and measured bulk RNAseq signal in the appropriate cell types (**Figure 3a,b**). We also found that our ATAC-RNA pairings and peak-to-gene links were sensitive to the cellular composition of our scRNA integration data. If the identical master peakset was compared to scRNA data from e10.5 to e11.5 mouse brain (“MOCA neuro”) or e9.5 to e13.5 mouse heart (“MOCA cardiac”)^74^ in place of our cMN-enriched scRNA data, we found fewer significant peak-to-gene links and fewer concordant cognate genes (**Figure 3c-f; Methods**).

**Figure 3.**
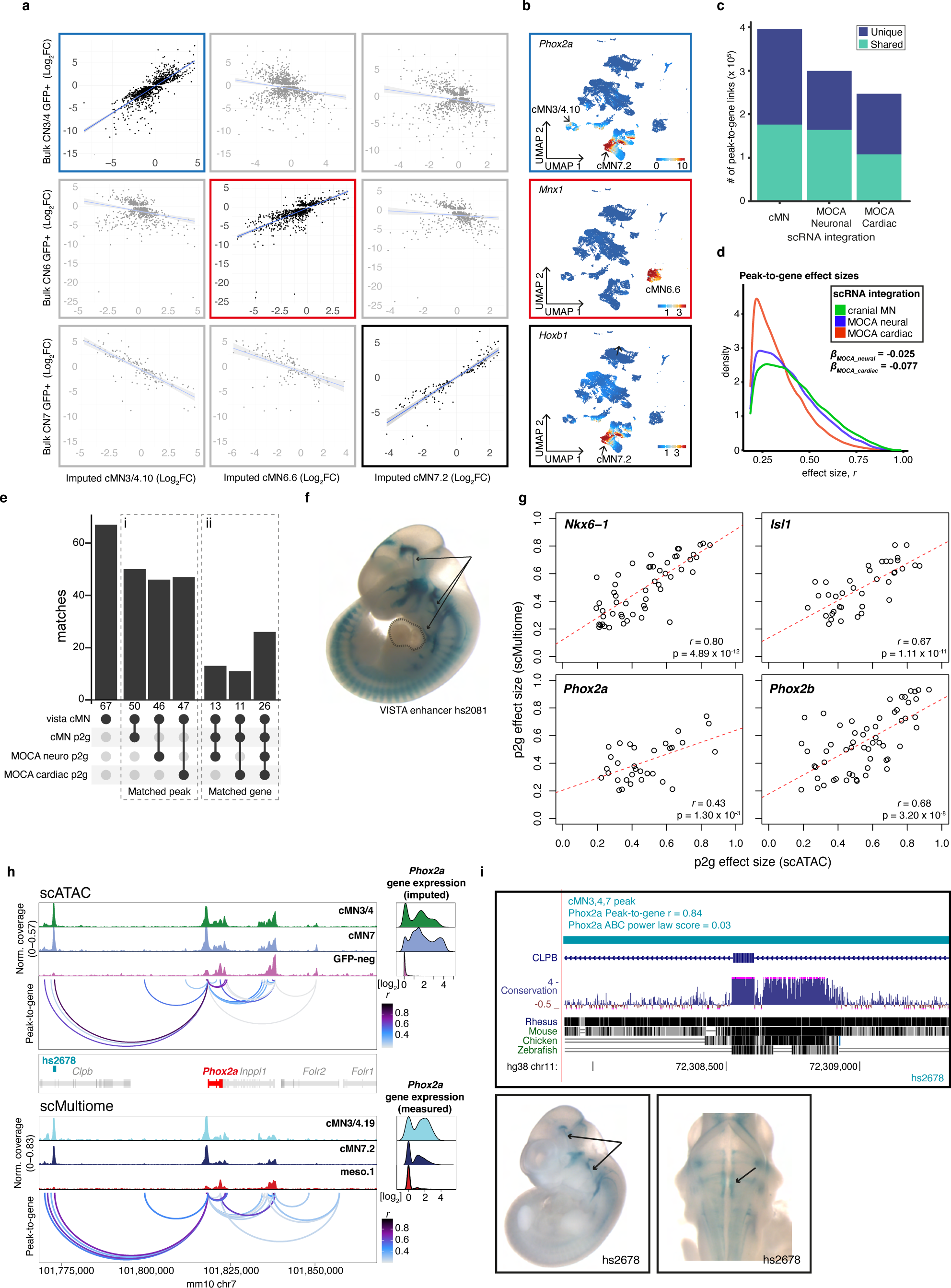
Effects of RNA input data on peak-to-gene accuracy. a. Scatterplots depicting imputed gene expression values projected onto scATAC clusters cMN3/4.10, cMN6.6, and cMN7.2 (x axis) versus measured gene expression values from independently collected bulk RNA-seq samples (y axis). Imputed gene expression shows a significant positive relationship when compared with corresponding bulk samples (cMN3/4, cMN6, and cMN7, respectively). b. Feature plots depicting imputed gene expression for three classic cMN marker genes (*Phox2a* (top, boxed in blue)*, Mnx1* (middle, boxed in red), and *Hoxb1* (bottom, boxed in black))^37^. Expression is restricted to corresponding clusters cMN3/4.10 (*Phox2a*), cMN6.6 (*Mnx1*), and cMN7.2 (*Phox2a, Hoxb1*) as expected. c. Stacked barplot depicting total number of unique and shared peak-to-gene links using three distinct scRNA integration datasets against the common scATAC cMN peakset. cMN: scRNA-seq data from age- and dissection-matched, oversampled cranial motor neurons (this work). MOCA Neuro: age-matched, uniformly sampled embryonic neural tissue from the MOCA database. MOCA Cardiac: non-age-matched, uniformly sampled embryonic cardiac tissue from the MOCA dataset^74^. d. Distribution of peak-to-gene effect sizes using different scRNA integration datasets (shared links only). Estimated effect sizes are significantly stronger using cMN scRNA integration when compared to MOCA neuro and MOCA cardiac integration. e. Barplot depicting peak-to-gene elements from the three scRNA integrations overlapping 67 experimentally validated cMN enhancers (“vista cMN”, left). i. “Matched peak” indicates overlapping peaks irrespective of predicted cognate gene (middle). ii. “Matched gene” indicates both overlapping peaks and identical cognate gene within the VISTA cMN enhancers (right, note that the vista cMN enhancers to not have defined target genes). Toggling between scRNA integrations can alter or eliminate target gene predictions. i and ii represent intersect and distinct peaks, respectively. f. *In vivo* enhancer assay for cMN VISTA enhancer hs2081 (lateral view). This enhancer overlaps a predicted peak-to-gene link using both cMN and MOCA cardiac scRNA input. However, enhancer activity is positive in cranial nerves 3, 7, and 12 (arrows) and negative in embryonic heart (dotted lines). g. Comparing scATAC versus scMultiome peak-to-gene effect sizes for four motor neuron transcription factors (*Nkx6-1*, *Isl1*, *Phox2a*, and *Phox2b*)^37^. Each circle represents a peak. All four genes show a positive linear relationship across both assays. h. scATAC (top) and scMultiome (bottom) accessibility profiles with peak-to-gene connections for a 100kb window centered around *Phox2a*. scATAC profiles are parsed by sample while scMultiome profiles are parsed by predicted cluster label. Peak-to-gene predictions are highly concordant across both assays. Novel cMN enhancer hs2678 is accessible in cMN3/4 and cMN7 and is predicted to enhance *Phox2a* by both scATAC (r = 0.84) and scMultiome (r = 0.69) peak-to-gene estimates. i. (Top) hs2678 orthologous region in the human genome. hs2678 is 70.3 kb distal to human *PHOX2A* and is embedded in coding and intronic sequence of *CLPB*. (Bottom) *In vivo* enhancer assay using human hs2678 sequence is positive in cMN3 and cMN7 (arrows), recapitulating known *Phox2a* gene expression patterns^41^. Reporter expression views are shown as lateral (left) and dorsal through the 4^th^ ventricle (right).

Next, we performed a joint ATAC-RNA coassay (“scMultiome”) on a subset of e11.5 GFP-positive cells represented in our main scATAC dataset (cMN3/4, cMN7, cMN12, sMN), thereby allowing us to benchmark our inferred ATAC-RNA pairings against direct experimental measurements (“vertical integration”; **Extended Data Figure 5a-d**). We found that scMultiome peak-to-gene links were highly concordant with our original scATAC peak-to-gene links (**Figure 3g-i**). We then examined the single cell accessibility profiles of four highly characterized cMN enhancers with known connection to the *Isl1* gene – a cMN master regulator embedded in a gene desert (**Figure 4a-c**)^58,75^. Strikingly, both by diagonal and vertical integration, we found that for these four enhancers (mm933, CREST1/hs1419, CREST3/hs215, and hs1321), chromatin accessibility alone was a significant predictor of *in vivo Isl1* expression patterns in the anatomically appropriate cMN (**Figure 4d,e**; **Extended Data Figure 5d**; Wald test p-value = 0.011; **Methods**).

**Figure 4.**
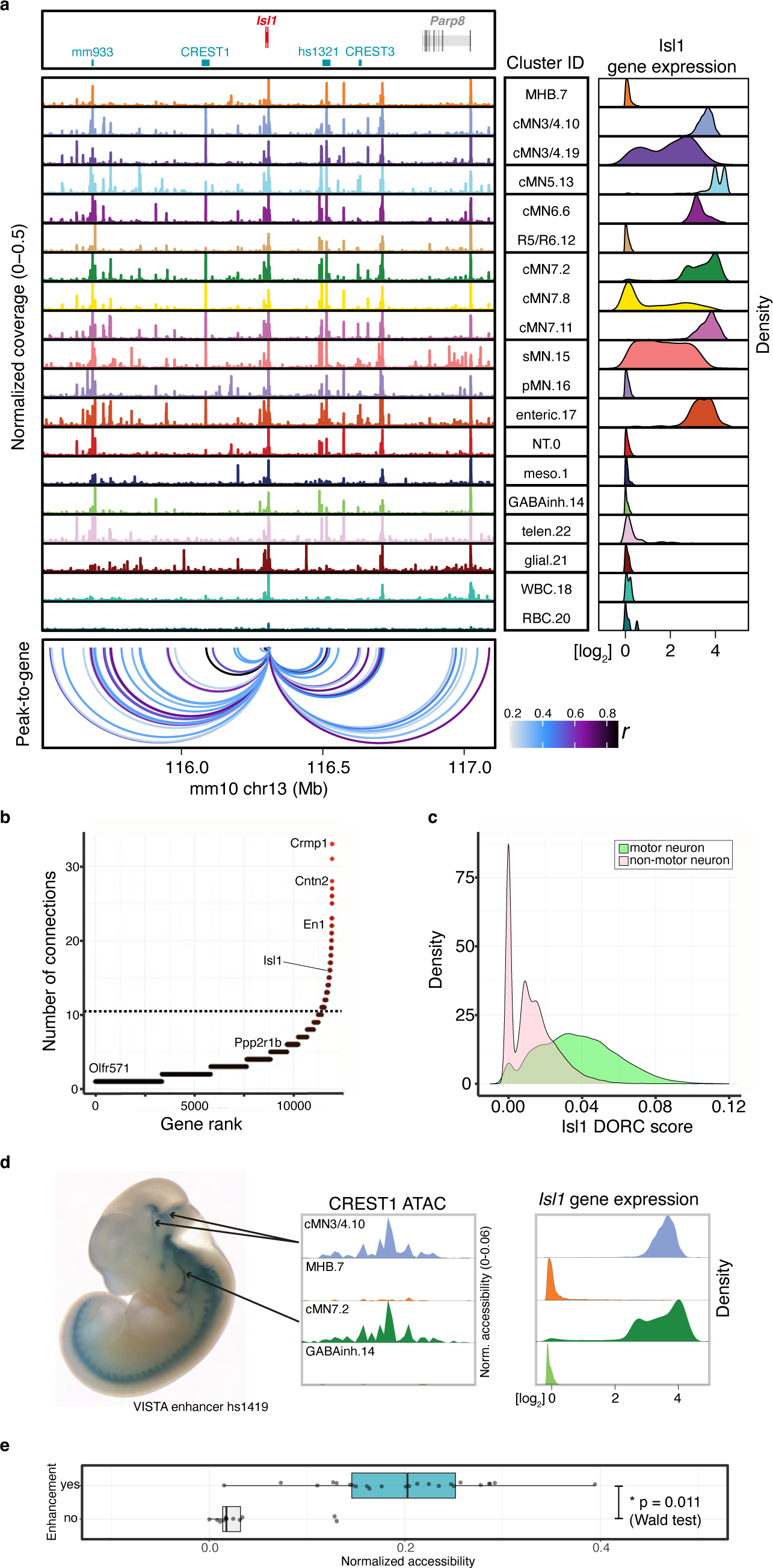
Exceptional gene regulation of cranial motor neuron master regulator *Isl1*. a. Pseudobulked chromatin accessibility profiles for all annotated clusters over a 1.5 Mb window centered about *Isl1.* Imputed gene expression profiles for each cluster are shown to the right. *Isl1* is located within a gene desert with the nearest up- and downstream flanking genes 1.2 and 0.7 Mb away, respectively. Peak-to-gene predictions match known *Isl1* enhancers (CREST1 in motor neurons and CREST3 in sensory neurons^75^; mm933 in multiple cranial motor nerves, dorsal root ganglion, and nose; hs1321 in multiple cranial motor nerves and forebrain) and identify additional putative enhancers surrounding *Isl1*. b. The number of normalized regulatory connections for each rank ordered gene. *Isl1* ranks in the top 1% of all genes with at least one regulatory connection. The inflection point of the plotted function is demarcated with a dotted line. c. Per-cell Domain of Regulatory Chromatin (DORC) scores for *Isl1* gene. DORC scores are significantly higher for cells from motor neuron clusters relative to non-motor neuron clusters (p-value < 1 x 10^-15^, ANOVA). d. (Left) Lateral whole mount *In vivo* reporter assay testing CREST1 (VISTA enhancer hs1419) enhancer activity. CREST1 drives expression in cranial nerves 3, 4, and 7 (black lines; there is also expression in trigeminal motor nerve). (Right) Single cell ATAC profiles and imputed gene expression for a subset of corresponding clusters. CREST1 accessibility and *Isl1* gene expression are positively correlated with *in vivo* expression patterns. e. Boxplot depicting normalized accessibility levels for enhancers CREST1, CREST3, mm933, and hs1321 within nine scATAC clusters corresponding to distinct anatomic regions. Manually scored enhancer activity (“enhancement”) is significantly correlated with normalized accessibility (p-value = 0.011, Wald test). Center line: median; box limits: upper and lower quartiles; whiskers – 1.5 x interquartile range.

Lastly, we integrated histone modification signatures into our enhancer predictions by performing H3K27Ac scCUT&Tag on e11.5 GFP-positive cMN3/4, cMN6, and cMN7 and e10.5 cMN7 (7 replicates total) and generated Activity-by-Contact (ABC) enhancer predictions for each cell type (**Methods**^76,77^). Of 6,072 total ABC enhancers, 4,925 (81%) directly overlapped our peak-to-gene links, including multiple *in vivo* ground truth enhancers (**Extended Data Figure 6a, Figure 3i, Figure 4a, Supplementary Table 7**). Because availability of cell type specific experimental data can be a limiting factor in accurate enhancer prediction, we assessed the relative contribution of cell type-specific chromatin accessibility versus histone modification data to ABC prediction accuracy. Specifically, among 67 annotated cMN enhancers in the VISTA enhancer database (visualized at e11.5 by presence of beta-galactosidase in the nucleus and/or nerve), 49 had some evidence of expression in cranial nerve (CN)7. Among these, we identified seven that had both visible CN7 expression and ABC cMN7 enhancer predictions at e11.5. For all seven enhancers (100%), ABC cognate gene predictions were concordant with peak-to-gene predictions. We then reran our ABC predictions, replacing either our cMN7 ATAC data with mouse embryonic limb e11.5 ATAC data (ENCODE ENCSR377YDY; “Limb ATAC”) or our cMN7 histone modification data with mouse limb histone modification data (ENCODE ENCSR897WBY; “Limb H3K27Ac”) and compared predictions. Substituting limb ATAC for cMN7 ATAC data resulted in only 14% (1/7) concordance, while substituting limb H3K27Ac for cMN7 H3K27Ac data resulted in 57% (4/7) concordance (**Extended Data Figure 6b**). Thus, for this curated set of data, we find that cell type-specific ATAC signal is a better predictor of reproducible cognate gene predictions than cell type-specific histone modification signal or non-cell-type-specific ATAC signal.

### Embryonic mouse chromatin accessibility atlas

In summary, we generated a chromatin accessibility atlas of the developing cMNs and surrounding cell types (reference tracks in the UCSC Genome Browser will be provided here). We combined GFP-positive (n = 49,708) and -negative (n = 36,381) cells to improve joint peak calling performance and to capture potential regional heterogeneity of non-motor neuron cell types as well as motor neuron progenitors^78^. Cluster analysis revealed 9 putative cMN, 4 putative sMN, and multiple non-MN/non-neuronal clusters (of 23 total). Although sMNs are not directly implicated in CCDDs, they may provide value for comparative studies with cMNs^79,80^. We also performed iterative clustering to identify 132 subclusters, of which 58 are highly pure groups of motor neurons. Although we are currently unable to annotate subclusters, more detailed spatial and developmental profiling of the cMN subnuclei may help to identify functionally-relevant groups of cells and/or cell states. Finally, a high quality and cell type-specific catalog of cMN elements and their cognate genes can be used to interpret and prioritize CCDD variants, as we describe below.

### Human phenotypes and genome sequencing

We enrolled and phenotyped 899 individuals (356 affected, 543 family members) across 270 pedigrees with CCDDs. 202 probands were sporadic (simplex) cases enrolled as trios, while 42 and 19 pedigrees displayed clear dominant or recessive inheritance patterns, respectively (**Supplementary Table 8**). Of note, the dominant pedigrees included 3 with CFP that we have reported to harbor pathogenic SNVs in a non-coding peak, “cRE2”, within the HCFP1 locus on chromosome 3^34^. The CCDDs included congenital fibrosis of the extraocular muscles (CFEOM), congenital ptosis (CP), Marcus Gunn jaw winking (MGJW), fourth nerve palsy (FNP), Duane retraction syndrome (DRS), congenital facial palsy (CFP), and Moebius syndrome (MBS) (**Supplementary Table 8**). Importantly, these CCDD phenotypes can be connected to maldevelopment of their disease-relevant cMNs: CFEOM to cMN3/4, CP to the superior branch of cMN3, FNP to cMN4, DRS to cMN6, CFP to cMN7, and MBS to cMNs 6 and 7 (**Figure 1a, Supplementary Table 1**). Affected individuals could have isolated or syndromic CCDDs.

We performed whole genome sequencing (WGS) and variant calling of the 899 individuals (**Methods**). First, to generate a comprehensive and unbiased set of genetically plausible candidates, we performed joint single nucleotide variant (SNV) and insertion/deletion (indel) genotyping, quality control, and variant frequency estimation from > 15,000 WGS reference samples in the Genome Aggregation Database (gnomAD)^81,82^. We identified 54,804,014 SNV/indels across the cohort. Of these, 1,150,021 (2.1%) were annotated as exonic, 18,761,202 (34.2%) intronic, 34,512,518 (63.0%) intergenic, and 364,300 (0.7%) within promoters. We next performed initial SNV/indel variant filtering based on established and custom criteria, including genotype quality, allele frequency, and conservation (**Methods**)^83,84^. We incorporated family structures to include or exclude genetically plausible candidates that are consistent with known modes of Mendelian inheritance. Applying this approach to the 54,804,014 SNVs/indels across our cohort, we identified 26,000 plausible candidates (mean = 101 variants per pedigree). We also performed short read structural variant (SV) discovery using an ensemble SV algorithm (GATK-SV) that was comparable to SVs generated in gnomAD and the 1000 Genomes Project^81,85^ and identified 221,857 total SVs (including transposable elements and other complex events). These WGS from deeply phenotyped CCDD pedigrees present a rich catalog of otherwise unannotated candidate Mendelian disease variants, as reflected in our report of noncoding SNVs and duplications as a cause of isolated facial weakness^34^.

### Integrating epigenomic filters with human WGS variants

To further refine the 26,000 CCDD candidate SNVs/indels, we eliminated from further analysis 37 pedigrees definitively solved by coding variants and reported separately, and then applied cell type-specific filters from our scATAC peakset to each CCDD phenotype (**Methods**). We identified 5,353 unique segregating SNVs/indels (3,163 *de novo/*dominant, 1,173 homozygous recessive, and 1,017 compound heterozygous) that overlapped cMN-relevant peaks of accessible chromatin (23.6 and 13.6 candidates per monoallelic and biallelic pedigree, respectively). Applying an analogous cell type-aware framework for SVs, we identified 115 candidates (72 deletions, 27 duplications, 1 inversion, 13 mobile element insertions, and 2 complex rearrangements encompassing multiple classes of SVs). There was substantial overlap between candidate variants and CCDD-relevant cMN peaks when compared to size-matched randomized peaks (median *de novo Z*-score = 10.9, median dominant inherited Z-score = 30.1, p-value < 2.0 x 10^-4^, permutation test; **Supplementary Table 9**). Using these 5,468 cell type-aware non-coding CCDD candidate SNVs/indels/SVs and ATAC-based cMN enhancers, we next identified strong candidate variants using gene-centric and peak-centric approaches.

We adopted a gene-centric aggregation approach by first identifying non-coding candidate variants connected to a restricted set of 16 known CCDD disease genes^19,21–26,28,42,86–93^. We identified non-coding variants connected to four: *MAFB*, *PHOX2A, CHN1,* and *EBF3* (**Table 1**). We also identified compound heterozygous variants connected to *ISL1* in a proband with CFP; *ISL1* is not a known disease gene but is a master cMN regulator (**Extended Data Figure 7a,b**). Extending this approach to the entire genome, we identified 559 genes with multiple connected peaks containing dominant candidate variants (“multi-hit genes”, range of connected variants per gene = 2-6, **Supplementary Table 10**).

**Table 1.** Non-coding candidate variants and putative target genes. _1_Coding loss-of-function intolerance -https://doi.org/10.1038/s41586-020-2308-7; _2_Coding dosage sensitivity -https://doi.org/10.1016/j.cell.2022.06.036; _3_Non-coding mutational constraint (1 kb windows) - https://doi.org/10.1101/2022.03.20.485034; _†_Multi-hit gene; _††_Multi-hit peak; _†††_non-coding deletion; *mean value across deleted interval; (Y) denotes established CCDD gene for stated phenotype; AD: autosomal dominant/de novo, ar(h): autosomal recessive homozygous, ar(ch): autosomal recessive compound heterozygous, I: intronic, P: promoter, D: distal, LoF: loss-of-function, GoF: gain-of-function.

| CCDD | Pedigree | Inheritance | Non-coding variant (hg38) | Peak Type | Nearest gene | Target gene | Distance to target (kb) | Reporter ID | Peak to gene r | Peak to gene FDR | gnomAD allele frequency | Predicted mechanism | SAD Z-score | Target gene loeuf<sup>1</sup> | Target gene pHaplo<sup>2</sup> | Target gene pTripl<sup>2</sup> | Non-coding Z-score<sup>3</sup> |
| --- | --- | --- | --- | --- | --- | --- | --- | --- | --- | --- | --- | --- | --- | --- | --- | --- | --- |
| CFEOM | S25 | AD | chr10:129794079 TTGAG>T | D | *EBF3* | *EBF3*<sup>†</sup> (Y-DRS) | 170 | hs2776 | 0.24 | 2.90E-07 | 8.37E-05 | LoF | -11.77 | 0.15 | 1.00 | 1.00 | 3.10 |
| MGJW | S176 | AD | chr10:129884231 C>A | I | *EBF3* | *EBF3*<sup>†</sup> (Y-DRS) | - | hs2775 | 0.29 | 3.89E-10 | 4.88E-05 | GoF | 0.11 | 0.15 | 1.00 | 1.00 | 3.74 |
| Ptois | S95 | AD | chr10:129944464 G>C | I | *EBF3* | *EBF3*<sup>†</sup> (Y-DRS) | - | hs2774 | 0.21 | 7.76E-06 | - | GoF | 0.98 | 0.15 | 1.00 | 1.00 | 5.14 |
| DRS | S12 | ar(h) | chr11:72394626 C>G | I | *CLPB* | *PHOX2A* (Y-CFEOM) | 156 | - | 0.26 | 1.09E-08 | 1.41E-03 | GoF | 0.18 | 0.80 | 0.76 | 0.98 | 2.32 |
| Ptois | S32 | AD | chr2:175005662 C>T<sup>††</sup> | P | *CHN1* | *CHN1* (Y-DRS) | - | - | 0.48 | 1.31E-28 | 1.39E-04 | LoF | -0.38 | 0.57 | 0.41 | 0.72 | 2.59 |
| CFEOM/DRS | S251 | AD | chr2:175006051 GCTT>G<sup>††</sup> | P | *CHN1* | *CHN1* (Y-DRS) | - | - | 0.48 | 1.31E-28 | - | GoF | 2.29 | 0.57 | 0.41 | 0.72 | 2.08 |
| DRS | S230 | AD | chr20:40866929-40945626<sup>†††</sup> | D | *TOP1* | *MAFB* (Y-DRS) | 256 | hs2769<br>hs2770 | 0.23* | 1.19E-05* | - | - | - | 0.40 | 0.94 | 1.00 | 2.19* |
| CFP | S205 | ar(ch) | chr5:51172762 T>A | D | *ISL1* | *ISL1* | 221 | hs1321 | 0.74 | 1.36E-86 | 2.26E-03 | LoF | -0.41 | 0.23 | 0.95 | 0.85 | -2.28 |
| CFP | S205 | ar(ch) | chr5:51172961 T>G | D | *ISL1* | *ISL1* | 221 | hs1321 | 0.74 | 1.36E-86 | 2.33E-03 | LoF | -0.12 | 0.23 | 0.95 | 0.85 | -2.28 |
| DRS | S190,<br>S238 | ar(h) | chr22:27493955-27497536<sup>††,†††</sup> | D | *MN1* | *MN1* | 307 | hs2757 | - | - | 1.38E-04 | - | - | 0.48 | 0.99 | 0.92 | 0.29* |
| DRS | S191 | ar(ch) | chr17:1455690 G>A<sup>††</sup> | I | *CRK* | *CRK* | - | - | - | - | - | GoF | 0.44 | 0.34 | 0.97 | 1.00 | 0.30 |
| DRS | S191 | ar(ch) | chr17:1456361 G>A<sup>††</sup> | P | *CRK* | *CRK* | - | - | - | - | 1.51E-03 | LoF | -1.24 | 0.34 | 0.97 | 1.00 | - |
| DRS | S211 | ar(ch) | chr17:1455565 C>T<sup>††</sup> | I | *CRK* | *CRK* | - | - | - | - | 1.19E-04 | GoF | 0.49 | 0.34 | 0.97 | 1.00 | 0.30 |
| DRS | S211 | ar(ch) | chr17:1456436G C>G<sup>††</sup> | P | *CRK* | *CRK* | - | - | - | - | 3.77E-04 | LoF | -12.28 | 0.34 | 0.97 | 1.00 | - |
| DRS | S211 | ar(ch) | chr17:1456438 G>A<sup>††</sup> | P | *CRK* | *CRK* | - | - | - | - | 3.77E-04 | LoF | -2.06 | 0.34 | 0.97 | 1.00 | - |
| DRS | WL | AD | chr17:48003752 A>C<sup>††</sup> | D | *CDK5RAP3* | *CDK5RAP3* | 22 | hs2777 | 0.57 | 8.04E-43 | - | GoF | 4.31 | 0.97 | 0.24 | 0.54 | 1.94 |
| MBS | S174 | ar(ch) | chr17:48003557 C>G<sup>††</sup> | D | *CDK5RAP3* | *CDK5RAP3* | 22 | hs2777 | 0.57 | 8.04E-43 | 4.04E-03 | LoF | -0.15 | 0.97 | 0.24 | 0.54 | 1.94 |
| MBS | S174 | ar(ch) | chr17:48003826 C>T<sup>††</sup> | D | *CDK5RAP3* | *CDK5RAP3* | 22 | hs2777 | 0.57 | 8.04E-43 | 9.42E-04 | GoF | 1.69 | 0.97 | 0.24 | 0.54 | 1.94 |
| CFP | S156 | AD | chr3:128459417G>C<sup>††</sup> | D | *DNAJB8* | *GATA2* | 7 | - | 0.28 | 6.08E-10 | - | LoF | -4.88 | 0.34 | 0.98 | 0.87 | - |
| CFP | S180 | AD | chr3:128459454A>G<sup>††</sup> | D | *DNAJB8* | *GATA2* | 7 | - | 0.28 | 6.08E-10 | 3.95E-05 | GoF | 2.88 | 0.34 | 0.98 | 0.87 | - |
| CFP | S194 | AD | chr3:128459455G>A<sup>††</sup> | D | *DNAJB8* | *GATA2* | 7 | - | 0.28 | 6.08E-10 | - | GoF | 11.40 | 0.34 | 0.98 | 0.87 | - |

*EBF3,* which encodes the EBF transcription factor 3, is an example of both a CCDD gene and a multi-hit gene. Monoallelic *EBF3* loss-of-function (LoF) coding mutations cause Hypotonia, Ataxia, and Delayed Development Syndrome (HADDS)^94^, and two individuals are reported with HADDS and DRS, one with a coding missense variant and one with a splice site variant^92,95^. We identified a series of coding and noncoding *EBF3* variants (**Supplementary Table 11**). Two probands with DRS have large *de novo* multi-gene deletions (**Figure 5a**), and one proband with fourth nerve palsy has a *de novo* stop-gain coding variant (**Figure 5b**). These three individuals also have phenotypes consistent with HADDS. We also identified three inherited non-coding variants with peak-to-gene connections to *EBF3* (**Figure 5b**). Pedigrees S25 (distal indel), S176 (intronic SNV), and S95 (intronic SNV) segregate non-coding candidate variants with isolated CFEOM, MGJW, and ptosis, respectively. The multiple ocular CCDD phenotypes we observed potentially reflect pleiotropic consequences of *EBF3* variants, a phenomenon previously observed for coding mutations in other CCDD genes^96^. Moreover, the differences in syndromic versus isolated phenotypes may reflect more cell type-specific effects of non-coding variants. Indeed, multiple Mendelian disorders with non-coding etiologies are restricted to isolated cell types or organ systems^57,65,97–100^. Notably, *EBF3* is broadly expressed across cMNs (**Figure 5c**) and is one of the most constrained genes in the human genome as measured by depletion of coding LoF variants in gnomAD and SV dosage sensitivity (loeuf = 0.1500 and pHaplo = 0.9996, respectively; **Figure 5d**)^82,101,102^. We observed exceptional conservation of non-coding elements within *EBF3* introns, comparable to or exceeding exonic conservation. This includes the ultraconserved element UCE318 (**Figure 5b,e**) located in intron 6 with a peak-to-gene link to *EBF3* (r = 0.69, FDR = 6.2 x 10^-69^). We also detected a peak-to-gene link from VISTA enhancer hs737 to *EBF3* (r = 0.60, FDR = 4.8 x 10^-49^), an element located > 1.2 Mb upstream of the gene that was previously reported to be linked to *EBF3* and to harbor *de novo* variants associated with autism with hypotonia and/or motor delay^103^. We did not observe any candidate variants in UCE318, consistent with extreme depletion of both disease-causing and polymorphic variation within ultraconserved elements^104^, nor in hs737, consistent with its non-CCDD phenotype.

**Figure 5.**
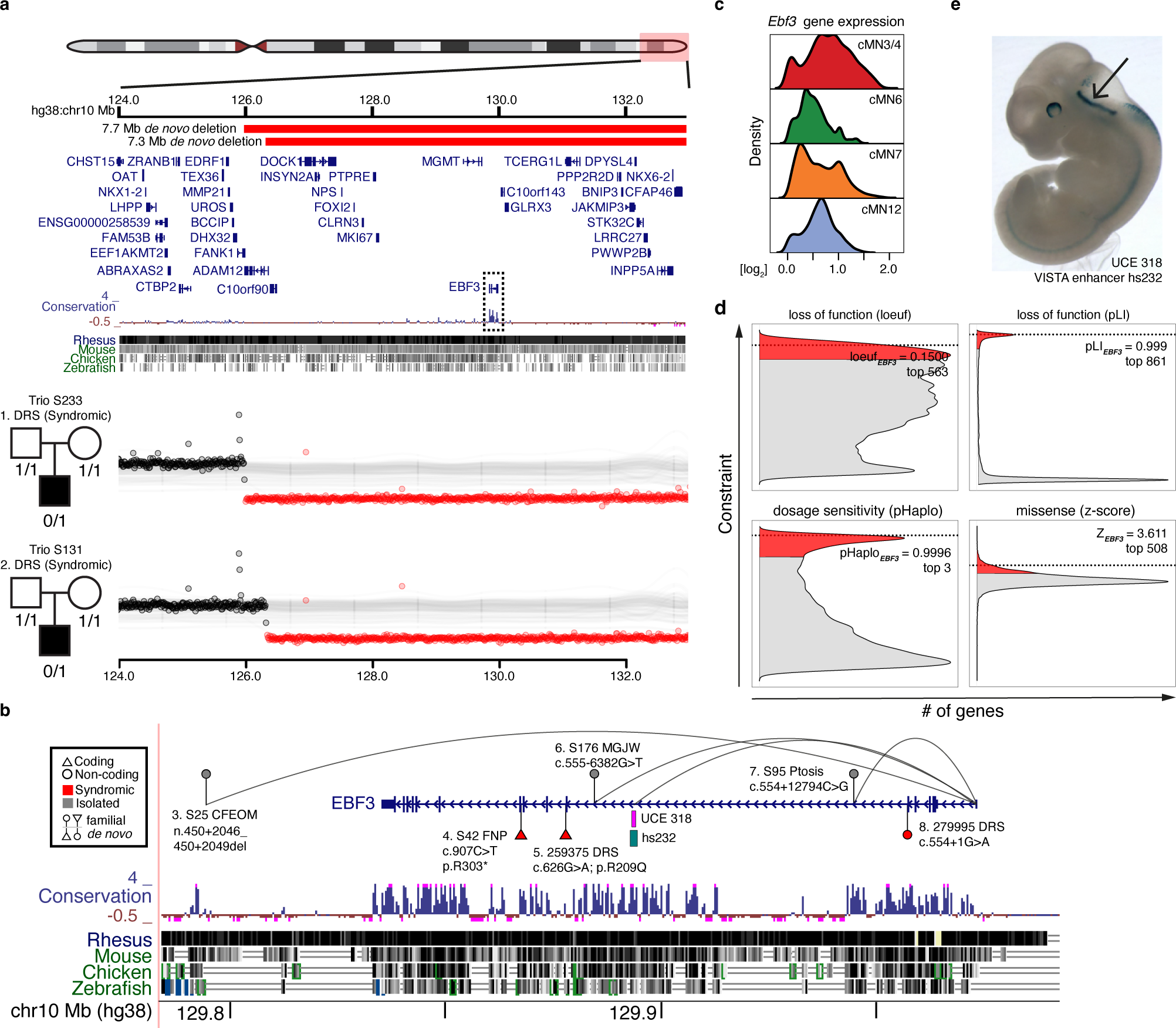
An integrated coding/non-coding candidate allelic series for *EBF3*. a. Window depicting the terminal arm of chr10q (top). Large *de novo* deletions in two trios (middle, bottom) with simplex syndromic DRS (S233, S131) overlap multiple coding genes including *EBF3* (boxed), an exceptionally conserved gene at the coding and non-coding level. b. Nominated coding and non-coding SNVs and indels connected to *EBF3*. For each variant, the subject’s WGS ID code, CCDD phenotype (and if isolated or syndromic), the variant coordinate in NG_030038.1 (and if coding or noncoding and if familial or *de novo)* is indicated. Variants 5 and 8 are reported previously in DECIPHER and elsewhere^92,95^. Peak-to-gene links containing variants connected to *EBF3* are depicted by curved lines. *EBF3* contains highly conserved non-coding intronic elements, including ultra-conserved element UCE 318 in intron 6, whose sequence drives strong expression in the embryonic hindbrain (VISTA enhancer hs232, see (e) below). c. Imputed gene expression profiles for *Ebf3*. *Ebf3* is broadly expressed among the cMNs. d. *EBF3* is exceptionally intolerant to loss-of-function, gene dosage, and missense variation. Density plots depict genome-wide distribution of loss-of-function constraint (“loeuf”, “pLI”)^82,125^, probability of haploinsufficiency (“pHaplo”)^101^, and missense constraint (“z-score”)^126^. Respective scores exceeding thresholds of 0.35, 0.9, 0.84, and 2.0 are colored red. *EBF3* (dotted lines) ranks as the 563^rd^, 861^st^, 3^rd^, and 508^th^ most constrained gene in the genome, respectively. Distributions are rescaled for consistent sign and ease of visualization. e. Lateral view of *in vivo* reporter assay testing UCE 318 (VISTA enhancer hs232), a putative *EBF3* enhancer (peak-to-gene r = 0.42, FDR = 6.72 x 10^-22^). Strong reporter expression is observed in the embryonic hindbrain (arrow).

Second, we took a peak-centric approach by examining all 5,468 (5,353 SNV/indels, 115 SVs) cell type aware non-coding variants, irrespective of cognate gene. When aggregating variants within appropriate cMN peak with corresponding CCDD phenotype, we identified 28 peaks harboring variants in more than one pedigree (“multi-hit peaks”). Fourteen multi-hit peaks contained variants obeying a dominant mode of inheritance (28 unique dominant/*de novo* variants with one variant present in two unrelated families, and including the 3 pathogenic chromosome 3 “cRE2” SNVs that cause CFP^34^), and 14 multi-hit peaks contained variants obeying a recessive mode of inheritance (35 unique recessive variants; **Supplementary Table 12**). Moreover, 10 of these multi-hit peaks were also linked to multi-hit genes. Because enhancers confer cell type-specific function, we reasoned that true functional non-coding SNV/indels are less likely than coding variants to cause syndromic, multi-system birth defects. Interestingly, when stratifying pedigrees by isolated/syndromic status, we found a significant overrepresentation of isolated CCDD phenotypes for our dominant multi-hit peaks (OR = 5.9, p-value = 2.3 x 10^-3^, Fisher’s exact test), but not for our recessive multi-hit peaks (OR 0.8, p-value = 0.64).

Among the multi-hit peaks, we identified 3.6 kb homozygous non-coding deletions centered over peak hs2757 in two probands with DRS; in each case, the consanguineous parents were heterozygous for the deletion. The probands had extended runs of homozygosity with a shared 16 kb haplotype surrounding the deletion, consistent with a founder mutation (**Figure 6a-c**). hs2757 is broadly accessible in multiple cMN populations, including cMN6, and is located 307 kb upstream of its nearest gene, *MN1*; *MN1* imputed gene expression estimates revealed widespread expression across all sampled cell types, including cMN6 (**Figure 6d**)^82,101^. Monoallelic LoF coding mutations in *MN1* cause CEBALID syndrome, a disorder affecting multiple organ systems. A subset of individuals with coding variants in *MN1* are reported to have CEBALID syndrome with DRS^89^. *MN1* is exceptionally constrained against LoF variation and dosage changes (loeuf = 0.087; pHaplo = 0.9901, **Figure 6e**)^82,101^ We performed *in vivo* enhancer testing on hs2757 which revealed reporter expression in a subset of tissues with known *Mn1* expression^105^, including expression in the hindbrain overlapping the anatomic territory of cMN6 (**Figure 6f**). Surprisingly, in this case we did not observe a peak-to-gene link between hs2757 and *Mn1* and did observe links with genes *C130026L21Rik* (whose sequence maps to a different chromosome in human) and *Pitpnb* (**Supplementary Table 12**). Multiple scenarios may explain this result, such as active *Mn1* enhancement occurring prior to the mouse e10.5-e11.5 window investigated here. Alternatively, our regression-based peak-to-gene estimates may be less sensitive at detecting enhancers for ubiquitously expressed genes, a phenomenon previously observed for other enhancer prediction methods^76^.

**Figure 6.**
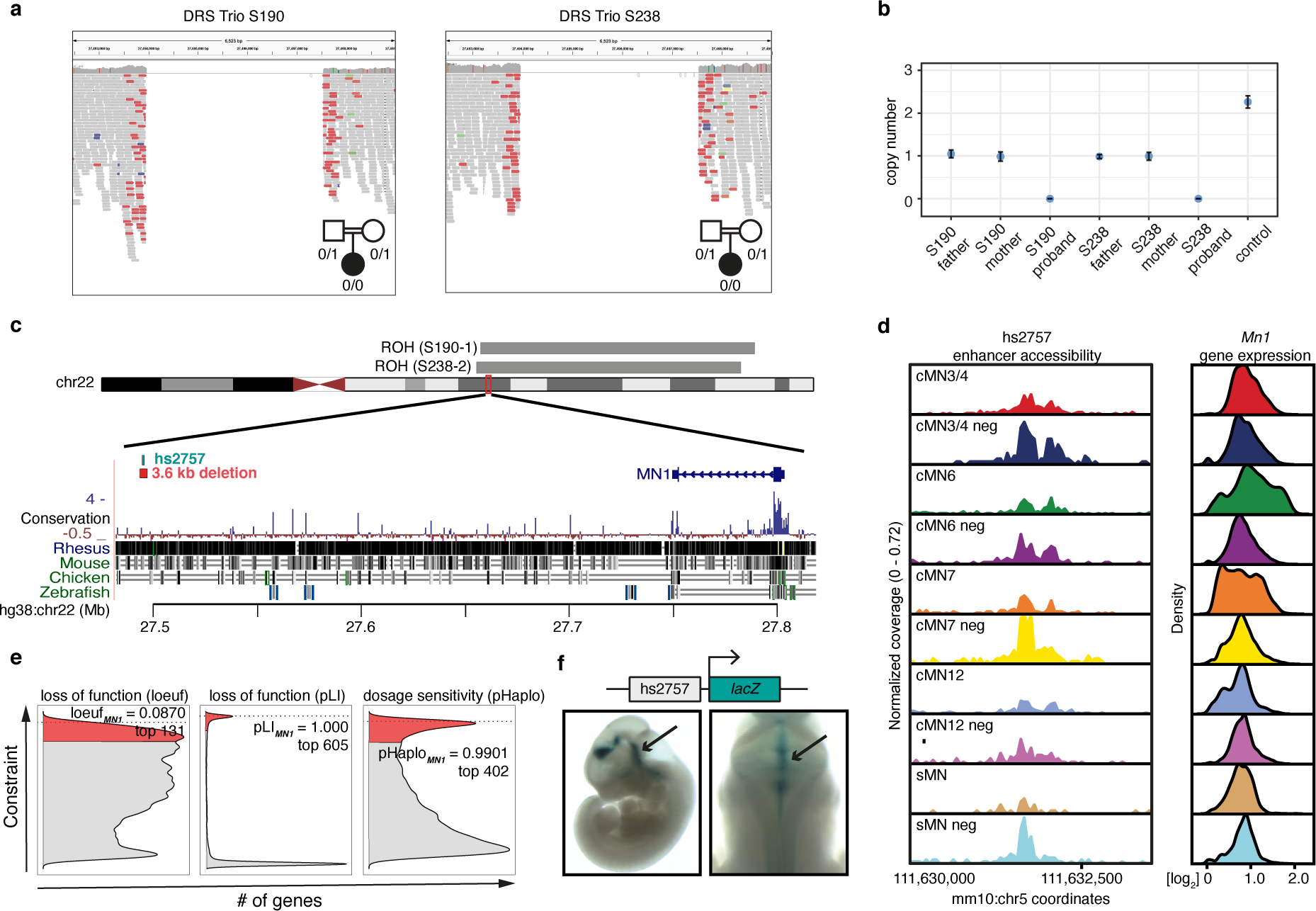
*MN1* enhancer deletions across multiple CCDD pedigrees. a. IGV screenshot depicting 3.6 kb non-coding deletions in two probands with DRS from separate consanguineous pedigrees (S190, S238). b. ddPCR copy number estimates of deletions. For each pedigree, the affected proband is homozygous recessive for the deletion with one heterozygous allele inherited from each parent. Error bars denote 95% confidence intervals. c. Genomic context of the non-coding deletions. The deletions (red bar below chr 22 ideogram) fall within extended runs of homozygosity (grey bars above ideogram, 19.5 Mb, 18.8 Mb, respectively, of which 16 kb surrounding the deletion is shared between the probands) and eliminates putative enhancer hs2757 (green bar below ideogram) located 307 kb from nearest gene *MN1*. d. hs2757 chromatin accessibility (left) and *Mn1* imputed gene expression (right) profiles in the cMNs and surrounding cell types. *Mn1* is widely expressed across multiple midbrain/hindbrain cell types, and hs2757 is accessible across multiple cell types, including cMN6. e. Density plots depicting genome-wide distribution of loss-of-function constraint (“loeuf”, “pLI”)^82,125^, and probability of haploinsufficiency (“pHaplo”)^101^ metrics. Respective scores exceeding thresholds of 0.35, 0.9, 0.84, and 2.0 are colored red. *MN1* (dotted lines) ranks as the 131^rd^, 605^th^, and 402^nd^ most constrained gene in the genome, respectively. Distributions are rescaled for consistent sign and ease of visualization. f. *In vivo* reporter assay testing hs2757 enhancer activity (humanized sequence). Lateral (left) and dorsal (right) whole mount *lacZ* staining reveals hs2757 consistently drives expression in midbrain and hindbrain tissue, including the anatomic territory of cMN6.

### Mechanistic insights of non-coding disease variants

Mendelian disease variant interpretation often relies on variant level predictions of pathogenicity^106,107^. However, such prediction algorithms are typically agnostic to cell type- or disease-specific information. More recent approaches have incorporated cell type-specific epigenomic data to annotate non-coding variants in common diseases^53,108,109^. To leverage our cell type-specific accessibility profiles for variant level functional interpretation, we trained a convolutional neural network^110^ to generate cell type-specific predictions of chromatin accessibility for each cranial motor neuron population. When evaluating held-out test data, we consistently observed high concordance between our accessibility predictions and true scATAC coverage for each cell type (median Pearson’s r = 0.84; range = 0.81 to 0.95; **Figure 7a**; **Extended Data Figure 8a-c**). Thus, to predict the effects of participant variants on element accessibility, we used our trained model to generate cell-type specific SNP Accessibility Difference (SAD)^110^ scores.

**Figure 7.**
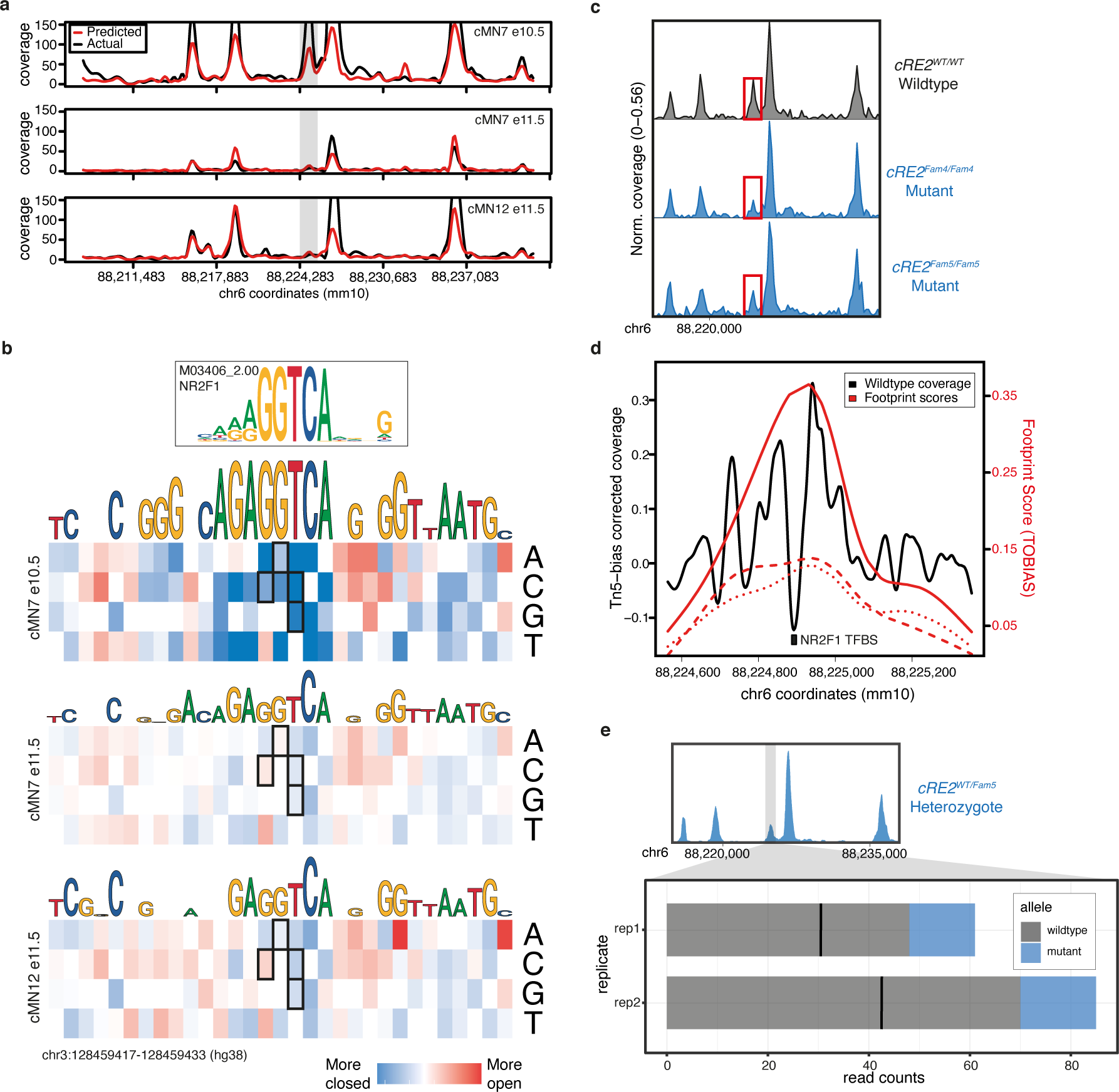
scATAC-trained convolutional neural network accurately predicts cell type specific accessibility status and human mutation effects in a transiently developing cell type. a. Neural net predicted chromatin accessibility profiles (red) compared to actual scATAC sequencing coverage (black) for a region of mouse chromosome 6 in three cell types (cMN7 e10.5, cMN7 e11.5, and cMN12 e11.5). The grey box highlights a transient 678 bp peak (cRE2) that is accessible in cMN7 e10.5, but not cMN7 e11.5 or cMN12 e11.5. SNVs within the human orthologous peak cRE2 cause congenital facial weakness, a disorder of cMN7. b. Neural net-trained *in silico* saturation mutagenesis predictions for specific nucleotide changes in human cRE2 for cMN7 e10.5, cMN7 e11.5, and cMN12 e11.5. Predicted loss-of-function nucleotide changes are colored in blue and gain-of-function in red. Predictions for four known loss-of-function pathogenic variants (chr3:128178260 G>C, chr3:128178261 G>A, chr3:128178262 T>C, chr3:128178262 T>G) are boxed. All four pathogenic variants are predicted loss-of-function for cMN7 e10.5, but not cMN7 e11.5 or cMN12 e11.5. c. Pseudobulk accessibility profiles of *cRE2* (red box) CN7 e10.5 for wildtype and two CRISPR-mutagenized mouse lines (*cRE2^Fam^*^4^*^/Fam^*^4^ *and cRE2^Fam^*^5^*^/Fam^*^5^) show a qualitative reduction in cRE2 scATAC sequencing coverage, consistent with *in silico* saturation mutagenesis predictions. Each pseudobulk profile represents normalized sequencing coverage across two biological replicates. d. Locus-specific footprinting evidence overlapping cRE2. A 792 bp window showing sequencing coverage for cMN7 e10.5 after correcting for Tn5 insertion bias. The NR2F1 transcription factor binding site is mutated in individuals with HCFP1-CFP and overlaps a local minimum in scATAC coverage. TOBIAS footprinting scores for *cRE2* wildtype, *cRE2^Fam^*^4^*^/Fam^*^4^, and *cRE2 ^Fam^*^5^*^/Fam^*^5^ are depicted in solid, dashed, and dotted lines, respectively. Wildtype footprinting scores are higher than mutant scores. e. Stacked barplot depicting wildtype versus mutant scATAC read counts over a 7.7 kb window for cMN7 e10.5 in *cRE2^WT/Fam^*^5^ heterozygote embryos. cRE2 mutant alleles are consistently depleted across two biological replicates (counts_WT_ / counts_MUTANT_ = 4.21; p-value = 2.4 x 10^-14^, binomial test).

Our peak-centric approach successfully re-identified the HCFP1 cRE2 SNVs that we reported to be pathogenic for CFP^34^, and scATAC data revealed that cRE2 was accessible in cMN7 at mouse e10.5 but not e11.5 (**Figure 7a**). Examining cRE2 SNV SAD scores, we found that all four Cluster A LoF variants were predicted to close the chromatin (SAD *Z*-scores of -4.88, -3.60, -6.29, and -3.93). Moreover, these predicted variant effects were specific to cMN7 at e10.5 (but not e11.5, **Figure 7b**), further underscoring the importance of accurately parsing both cell type and developmental cell state. We then experimentally corroborated the predicted variant effect on chromatin accessibility by performing scATAC on two CRISPR-mutagenized mouse lines harboring HCFP1 cRE2 Cluster A SNVs (previously reported *cRE2^Fam^*^5^*^/Fam^*^5^ and new *cRE2^Fam^*^4^*^/Fam^*^4^ mouse models)^34^. Consistent with our machine learning predictions, we observed subtle yet consistent reductions in *cis* chromatin accessibility for both mutant lines when compared to wildtype (4/4 replicates total; mean normalized mutant / wildtype coverage = 0.59; **Figure 7c**). We also found positive evidence for site-specific footprinting overlapping the cRE2 NR2F1 binding site in wildtype, but not in the two mutant lines (**Figure 7b,d**), consistent with results from targeted antibody-based assays^34^. Finally, to circumvent batch and normalization effects across separate experiments, we performed scATAC on embryos from wildtype-by-mutant crosses from *cRE2^Fam^*^5^*^/Fam^*^5^ and directly measured the resultant heterozygous mutant allele fraction in *cis* (“binomial ATAC”; **Figure 7e**). This approach generates an internally calibrated estimate of effect size and is sufficiently powered to detect true differences at relatively low sequencing coverage (i.e., chromatin accessibility profiles of rare or transiently developing cell types). We found a significant depletion of *Fam5* mutant alleles across multiple replicates, again consistent with a LoF mode of pathogenicity (wildtype / mutant counts = 4.2; p-value = 2.4 x 10^-14^; binomial test). These multiple lines of evidence, both at the epigenome-wide level and at a well-characterized individual locus provide support that our machine learning model is well calibrated and not overfitted.

We next examined the predictions of the neural net at epigenome-wide level, and among our 5,353 cell type-aware candidate SNVs/indels, identified 114 additional variants with normalized absolute SAD *Z*-scores > 2; that is, variants predicted to significantly increase or decrease accessibility in *cis* within their disease-relevant cellular context, including 7 variants linked to multi-hit genes (**Supplementary Table 13**). When incorporating these SAD scores, we identified several cell type-aware candidate variants and peaks with convergent lines of evidence. First, several of the non-coding variants connected to known CCDD genes had significant SAD scores (**Table 1**). The EBF3 non-coding variants chr10:129794079TTGAG>T, chr10:129884231C>A, and chr10:129944464G>C had SAD scores of -11.77, +0.11, and +0.98, respectively. The variant connected to *CHN1* segregated in a parent and child with a mixed CFEOM-DRS phenotype was predicted to increase accessibility (SAD Z-score = +2.29). This is notable because *CHN1* coding variants result in atypical DRS through a gain-of-function mechanism^23,43,111^. Second, combining multiple layers of evidence can be used to elevate candidate variants connected to potentially novel CCDD disease genes. For example, compound heterozygous variants in two DRS probands in the multi-hit *CRK* promoter region had significant negative scores consistent with LoF (SAD Z-scores = -13.69, -2.06; **Supplementary Table 12**). Such highly annotated non-coding variants are attractive candidates for downstream functional validation, as they provide distinct, refutable predictions for gene targets, cell types, and effect on accessibility.

### Nominated cell type-specific variants alter expression *in vivo*

Although we show that single cell chromatin accessibility is a strong predictor of cMN enhancer activity, even highly conserved and presumably functional enhancers can be surprisingly robust to mutagenesis^8,112–114^. Therefore, to evaluate the functional consequences of our nominated CCDD variants, we selected 33 elements harboring cell type-aware candidate SNVs for *in vivo* humanized enhancer assays. For testing, we prioritized these variants based on multiple annotations from our framework, including conservation, significant SAD scores, multi-hit peaks/genes, and cognate gene predictions (**Supplementary Table 14**). We first screened the wildtype human enhancer sequences and detected positive enhancer activity in 82% (27/33) of candidates. Combining these with the 26 previously tested, we found enhancer activity in 44/59 total (75%). Importantly, we note that these elements were not selected randomly and therefore not intended to reflect generalizable patterns across the genome.

Next, we tested 4 of the 27 positive elements by introducing the nominated CCDD SNVs into the wildtype sequence. Remarkably, one mutant enhancer harboring multiple candidate variants for DRS and MBS (“hs2777-mut”) showed visible gain of expression compared to wildtype (“hs2777”), including in midbrain, hindbrain, and neural tube (**Extended Data Figure 9a,b**). Wildtype hs2777 is accessible across multiple cell types and has peak-to-gene links to seven genes (*Cdk5rap3, Nfe2l1, Sp2, Tbx21, Npepps, Socs7*, and *Snx11*), and ABC enhancer prediction for *Cdk5rap3*, specifically to cMN7 at e10.5. hs2777-mut contains four SNVs (1 DRS, 2 MBS, 1 off-target, mutating 0.21% of original wildtype base pairs; **Extended Data Figure 9c,d**). To better decompose the individual effects of these variants, we performed *in silico* saturation mutagenesis across the entire hs2777 sequence (**Extended Data Figure 9e**). We observed notable gain-of-function effects for two of the three on-target SNVs (DRS “Variant C”, and MBS “Variant D”; chr17:48003826C>T and chr17:48003752A>C) within the affected cell types, with corresponding SAD Z-scores ranging from +1.12 to +4.34.

## DISCUSSION

We have developed a publicly available atlas of developing cranial motor neuron chromatin accessibility and have combined it with cell type-specific histone modification and *in vivo* transgenesis information to generate a reference set of enhancers with cognate gene predictions in a set of rare, transiently developing cell types. Such a resource can be used to discover highly specific cREs and target genes underlying the molecular regulatory logic of cMN development. Furthermore, we can leverage known properties of the cMNs to inform comparative studies across diverse cell types. For example, the ocular cMNs are known to be selectively resistant to degeneration (compared to sMNs) in diseases such as ALS. Therefore, understanding the differentially accessible cREs that underlie differences between cMNs/sMNs could render important clues to the mechanisms of selective resistance/vulnerability and ultimately open new therapeutic avenues^80^. Finally, a deeply sampled, highly specific chromatin accessibility atlas may help to learn generalizable features that predict enhancer activity in additional cell types. Importantly, cranial nerve expression is a core readout for tested cREs in the VISTA enhancer database, thereby providing invaluable ground truth data at an overlapping developmental timepoint (e11.5)^58^.

We used this reference to nominate and prioritize non-coding variants in the CCDDs, a set of Mendelian disorders altering cMN development and demonstrate that principled prioritization approaches can select appropriate candidates for downstream functional validation (e.g., transgenic reporter assays, non-coding *in vivo* disease models, etc.), which are otherwise often costly and labor-intensive with high rates of failure. To aid in interpretation, we connected non-coding variants to their cognate genes using imputed gene expression values from separate assays (diagonal integration). This approach allowed us to leverage existing information of cognate coding genes, including known disease associations and coding constraint^82^. Moreover, such integrated cell type-aware datasets provide important context to cell type-agnostic estimates of non-coding constraint (discussed in ref. ^115^). When applying this framework to our CCDD cohort, we achieved a search space reduction of 4 orders of magnitude, making non-coding candidate sets human-readable and tractable for functional and mechanistic studies (23.6 candidates per monoallelic pedigree; 13.6 per biallelic pedigree). Furthermore, we incorporated multiple lines of evidence such as allelic aggregation, cognate gene identification, mutational constraint, and functional prediction. This approach successfully re-identified the pathogenic variants in our cohort at the *GATA2* cRE2 locus^34^ and led us to nominate novel candidate disease variants (**Table 1**). We also identified compelling individual candidate variants and peaks without multiple hits. Such candidates will be easier to resolve with larger cohort sizes and larger families. Indeed, our ability to reduce candidate variant numbers was limited by the large proportion of unsolved small dominant pedigrees in our cohort, which are notoriously difficult to analyze. Moreover, while *de* novo and recessive mutations are clearly an important source of causal pathogenic variation in sporadic cases, such cases are also more likely to involve non-genetic etiologies.

Although a given peak can harbor hundreds of predicted transcription factor binding motifs, we demonstrate in principle that locus-specific footprinting can implicitly reduce a ∼1 kb peak to a ∼10 bp individual transcription factor binding site of interest. Given sufficient sequencing coverage^116^ and data quality, such approaches could immediately be applied to other rare diseases and cell types. Alternatively for common diseases, causal non-coding variants are more abundant, but also confounded by linkage disequilibrium. In this case, locus-specific footprinting (in concert with careful demarcation of element boundaries, chromatin accessibility QTL analysis^117^, and statistical fine-mapping^118^) may further resolve causal common variants and identify affected transcription factor binding sites across the genome – all inferred from a single assay. Proof of feasibility of such approaches in rare diseases could also influence data collection strategies for common diseases^119^.

Through our analysis, we also encountered potential limitations affecting non-coding variant interpretation. We in part leveraged sequence conservation and constraint to prioritize pathogenic variants. However, while the known genes and cREs underlying cMN development are highly conserved, a conservation-based strategy may not identify pathogenic variants in human-specific and/or rapidly evolving sequences^114,120,121^. Strikingly, we also found that even relatively subtle differences in cellular composition and ATAC/RNA collection strategies can distort cognate gene estimates. These findings should inform appropriate sampling strategies in the future, such as single cell multiomic assays. Unbiased genetic strategies such as partitioned LD score regression can be extremely useful towards defining disease-relevant cell types, though such approaches are effectively restricted to common diseases^122^. Moreover, we find that even when sampling the appropriate cell type, subtle differences in cell state can profoundly influence variant interpretation. We provide a concrete example at the well-characterized non-coding *GATA2* locus^34^, where pathogenic variant effects are no longer detectable in the same cell type within a mere 24 hours of development (i.e., embryonic day 10.5 versus 11.5). Moreover, we sampled cMNs at e10.5 and e11.5 based on developmental patterns of previously described protein-coding mutations, but we do not exclude the possibility that novel disease mutations may also be relevant at different timepoints. Therefore, while our genetic framework can generalize to other disorders, we suspect that appropriate prospective or retrospective epigenomic cell sampling will benefit from highly detailed biological knowledge of each specific disease process.

Finally, the interpretation of non-coding variants can benefit from our knowledge of coding variants as they share challenges in common – namely, practical limitations in allelic expansion and functional validation. Here, we present generalizable approaches that aggregate plausible alleles based on physical (“peak-centric”) and biological (“gene-centric”) proximity to facilitate allelic expansion in a principled manner. These challenges may be further alleviated by expanding rare disease data sharing platforms^123^ to more comprehensively incorporate non-coding variation. Finally, development of functional perturbation assays that balance both scalability^113^ and specificity^124^ will disproportionately benefit validation of non-coding variants, which are naturally more abundant and cell type-specific than coding variants. The outputs of such assays would also iteratively provide training material for further refined functional prediction algorithms.

Rapid advances in next generation sequencing technologies have led to a renaissance in Mendelian gene discovery. As access to WGS and functional genomics data becomes less limiting, alternative analytical and experimental frameworks will be needed to finally resolve Mendelian cases and disorders that are otherwise recalcitrant to traditional exome-based approaches.

## ACKNOWLEDGEMENTS

We are indebted to all study participants and their families. We thank Ryosuke Fujiki, Tulsi Patel, Ben Weisburd, Julie Jurgens, Orit Rozenblatt-Rozen, Aviv Regev, Andrew Hill, and Jay Shendure for important technical discussions. We thank Max Tischfield, Sarah Izen, Alicia Nugent, Alon Gelber, and Matthew Bauer for technical assistance with bulk and scRNA-seq experiments. Next generation sequencing for single cell experiments was performed at the Molecular Genetics Core at Boston Children’s Hospital. WGS of the CCDD cohort was performed at Baylor College of Medicine through the Gabriella Miller Kids First Pediatric Research Program (dbGaP Study Accession: phs001247). New mouse lines were generated by the Gene Manipulation & Genome Editing Core at Boston Children’s Hospital. FACS experiments were performed at the Blavatnik Institute Department of Immunology Flow Cytometry Core Facility at Harvard Medical School, the Boston Children’s Hospital Hem/Onc-HSCI Flow Cytometry Research Facility, and the Dana-Farber Flow Cytometry Hematologic Neoplasia and Jimmy Fund Cores at Dana-Farber Cancer Institute.

The work was supported by the Gabriella Miller Kids First Pediatric Research Program NHBLI X01HL132377 (E.C.E.), NEI R01EY027421 (D.G.M., M.E.T., E.C.E.), NICHD R01HD114353 (L.A.P), NHGRI R01HG003988 (L.A.P.), NIMH R01MH115957 (M.E.T., H.B.), DP2-AG072437 (E.A.L.), NINDS K08-NS099502 (M.F.R.), NHLBI T32-HL007627 (M.F.R), NIGMS T32-GM007748 (M.F.R.), Project ALS A13-0416 (E.C.E.), Boston Children’s Hospital - Broad Institute Collaborative Grant (E.C.E.), Boston Children’s Hospital Manton Center Rare Disease Fellowships (A.S.L, B.Z.) and Manton Center Pilot Project Award (B.Z.), Suh Kyungbae Foundation (E.A.L.), the Abramson Fund for Undergraduate Research (C.L.), and the Boston Children’s Hospital Intellectual and Developmental Disabilities Research Center (NIH U54HD090255). The research of M.K. and L.A.P. was conducted at the E.O. Lawrence Berkeley National Laboratory and performed under U.S. Department of Energy Contract DE-AC02-05CH11231, University of California. E.C.E. is an Investigator of the Howard Hughes Medical Institute.

## CONTRIBUTIONS

A.S.L. and E.C.E. led the experimental design. A.S.L., L.J.A., M.K., W.-M.C., B.P., M.F.R., and A.P.T. performed experiments. A.S.L. led the computational analysis. A.S.L., L.J.A., L.N.F., T.E.C., B.Z., A.S-J., J.M.F., I.W., X.Z., C.L., K.M.L., M.L., and H.B. performed computational analysis. A.S.L., W.-M.C., B.J.B., V.R., and E.C.E processed human samples and data. D.G.M., E.A.L., M.E.T., H.B., L.A.P., and E.C.E. provided funding and project supervision. A.S.L. and E.C.E. wrote the manuscript. A.S.L. devised the study. E.C.E. oversaw the study. All authors read and approved the manuscript.

## FIGURE LEGENDS

**Extended Data Figure 1.**
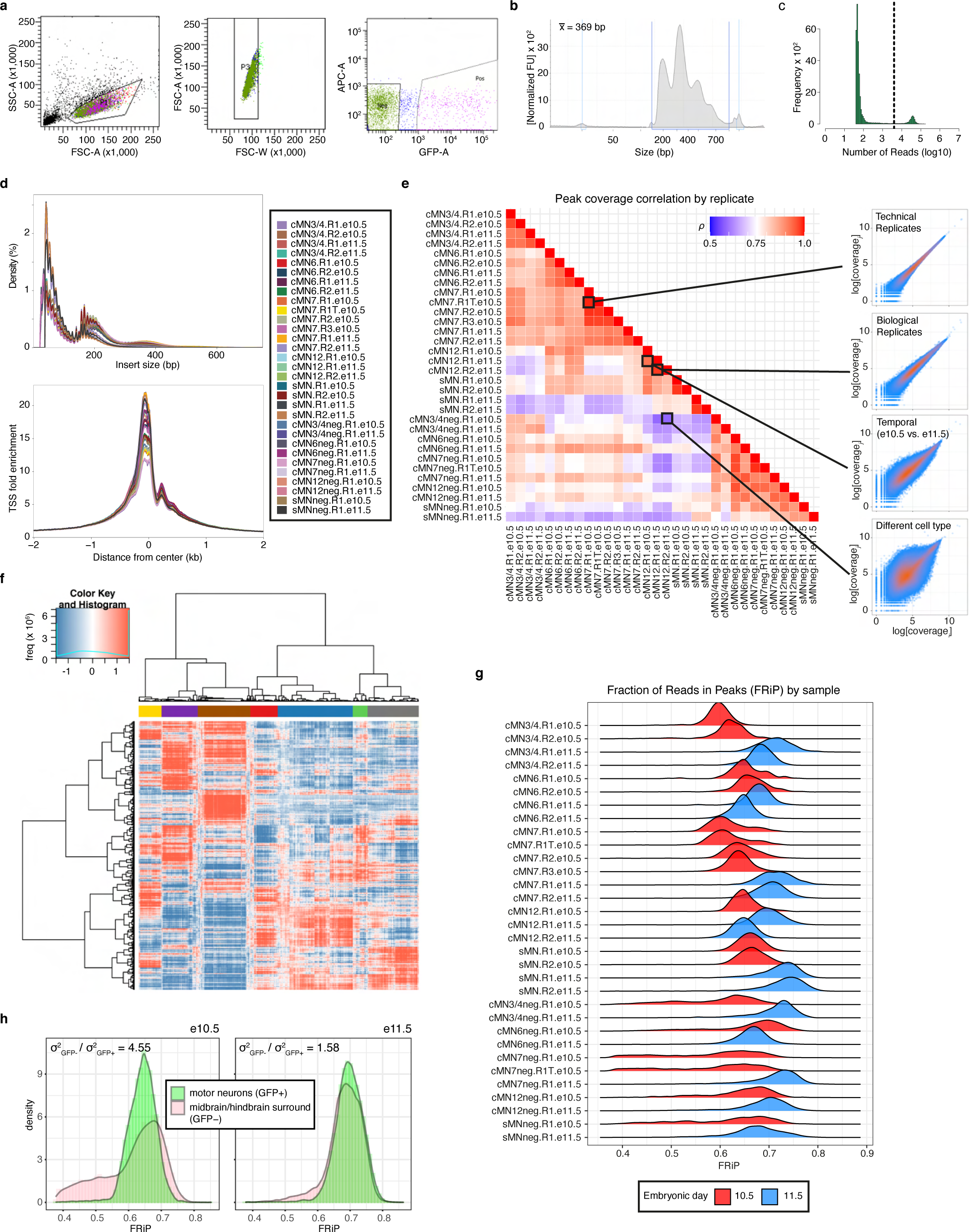
Per-cell and -sample quality metrics for scATAC data. a. Representative FACS gating strategy for WT GFP-positive and GFP-negative cMN7 at e10.5. Left: Forward scatter area (FSC-A) and side scatter area (SSC-A), corresponding to cell size and granularity/complexity, are used to enrich for intact cells and exclude debris. Middle: forward scatter width (FSC-W) and FSC-A are used to exclude doublets. Right: Green fluorescent protein area (GFP-A) and 633 nm-excitation (APC-A) are used to enrich for GFP-positive and GFP-negative cells. GFP-negative gates are calibrated by dissociated limb buds prior to collection as a negative control. All samples are fresh, live cells without fixative or nuclear staining. b. Representative TapeStation trace showing tagmented DNA fragment sizes prior to library preparation. c. Representative histogram of per-cell scATAC reads in a single sample. Read cutoff is shown by a dotted line and determined heuristically for each sample. d. Insert size distributions (top) and transcriptional start site (TSS) enrichment (bottom) for all samples and replicates. Insert sizes consistently show a characteristic nucleosome banding pattern (∼147 bp wavelength). Samples IDs are shown in **Supplementary Table 2**. e. Correlation matrix depicting all possible pairwise sample correlations (Spearman’s rho) for scATAC coverage in all rank-ordered peaks. Scatterplots for selected sample pairs from the four highlighted boxes within the matrix are shown on the right. Correlations decrease with increasing biological distance (top to bottom). f. Representative clade diagram depicting the relative accessibility (red is positive, blue is negative) of 5kb genomic windows (rows) across individual cells within a given sample (columns). Distinct clades (colored bars) were determined heuristically for each sample for downstream peak calling. The number of clades per sample were selected to maximize representation of common and rare cell types. g. Ridgeplot depicting density of per-cell fraction of reads in peaks (FRiP) for each dissected sample and replicate at e10.5 (red) and e11.5 (blue). Samples IDs are shown in **Supplementary Table 2**. Mean FRiP values are consistently higher for e11.5 samples (p-value = 4 x 10^-5^, binomial test). h. Distribution of FRiP values for GFP-positive motor neurons (green) versus GFP-negative surrounding brain tissue (pink). GFP-negative cells display significantly greater dispersion compared to GFP-positive cells, particularly at e10.5. (p-value = 1.1×10^-286^, Brown-Forsythe Test). See **Supplementary Note 1** for additional information.

**Extended Data Figure 2.**
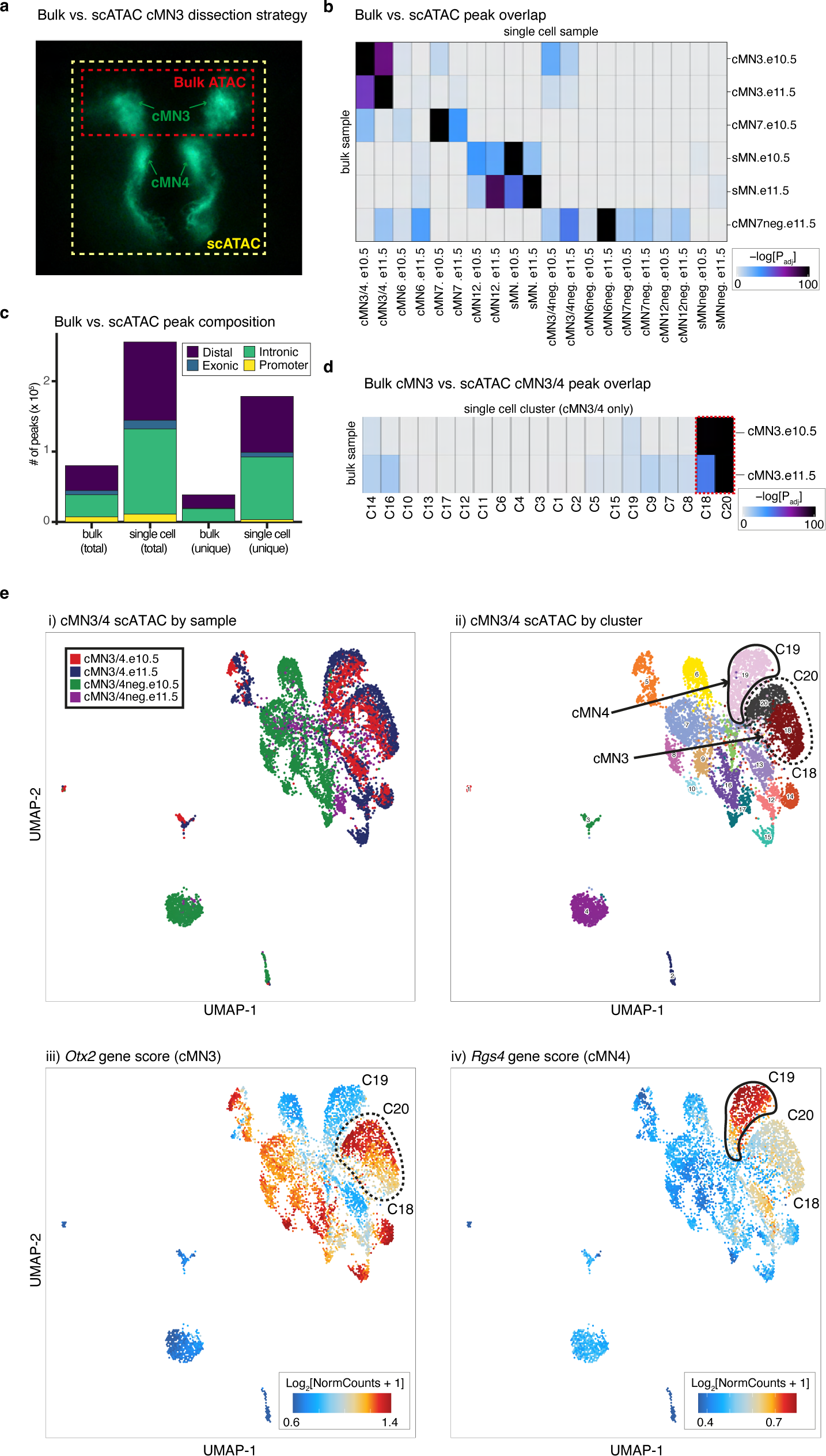
Comparing and contrasting bulk versus single cell ATAC profiles. a. Fluorescence microscopy image illustrating cMN3 and cMN4 microdissection strategies. For scATAC experiments, cMN3 and cMN4 were microdissected *en bloc* (yellow box). For bulk ATAC microdissections, only cMN3 was excised (red box). All other cMN microdissection strategies were identical across bulk and scATAC. b. Heatmap depicting enrichment of sample-specific bulk ATAC versus scATAC peaks. Color scale represents hypergeometric test p-values using the *peakAnnoEnrichment()* function in *ArchR*. Samples marked with “neg” are GFP-negative cells surrounding the motor neurons of interest. All other samples are GFP-positive motor neurons. c. Stacked barplot depicting relative proportions of different classes of accessible chromatin (“distal”, “exonic”, “intronic”, and “promoter”). scATAC peaks are enriched for total number of peaks, total number of unique peaks, and cell type-specific peak annotations (distal and intronic). d. Heatmap depicting enrichment of overlapping peaks for bulk cMN3 dissections versus *ad hoc* clusters (C1-C20) generated from scATAC cMN3/4 dissections only. Color scale represents hypergeometric test p-values*. Ad hoc* clusters C18 and C20 with the highest peak enrichment for bulk cMN3 are outlined by dashed red lines. e. *In silico* microdissection of scATAC cMN3/4 clusters corroborates physical microdissections. Left to right, UMAP embeddings of scATAC cMN3/4 dissections colored by i) dissected sample; ii) *ad hoc* clusters; and gene scores for iii) cMN3 marker gene *Otx2*^126^; and iv) cMN4 marker gene *Rgs4*^127^. Putative cMN3 (C18 and C20) and cMN4 (C19) clusters inferred from dissection origin, marker genes, and GFP status are denoted by dashed and solid red lines, respectively.

**Extended Data Figure 3.**
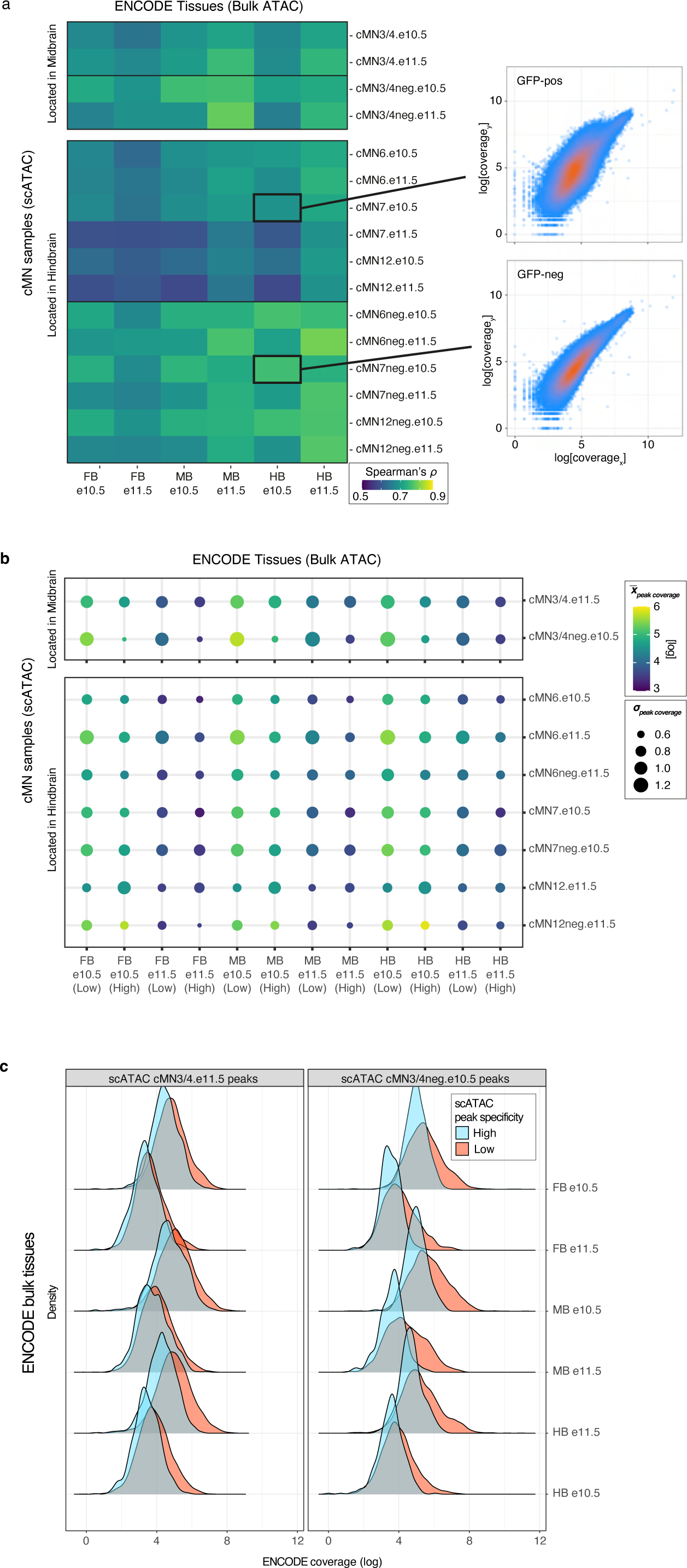
Cranial motor neuron scATAC peaks are underrepresented in regional bulk datasets. a. (Left) Heatmap depicting correlation coefficients (Spearman’s *ρ*) between scATAC peaks from cMN microdissections versus bulk ATAC peaks from ENCODE e10.5 and e11.5 mouse developing forebrain (FB), midbrain (MB), and hindbrain (HB) dissections. Anatomically concordant bulk brain regions are more highly correlated with scATAC non-motor neuron samples (‘-neg’) than scATAC cranial motor neuron samples. (Right) Scatterplots depicting rank-ordered per-peak sequencing coverage for bulk vs. scATAC samples. b. Bubble chart depicting ENCODE bulk ATAC coverage in scATAC cMN peaks from a subset of samples, stratified by cell type specificity scores (‘High’ vs. ‘Low’). Colors reflect mean peak coverage (with lighter color reflecting higher coverage), while area reflects standard deviation. Bulk tissues tend to have higher coverage in low specificity peaks when compared to highly cell type specific peaks. c. Density plots depicting distribution of ENCODE bulk peak coverage within cMN3/4 scATAC peaks from (b), stratified by specificity scores. High specificity scATAC peaks (blue) have consistently lower bulk coverage compared to low specificity peaks (red).

**Extended Data Figure 4.**
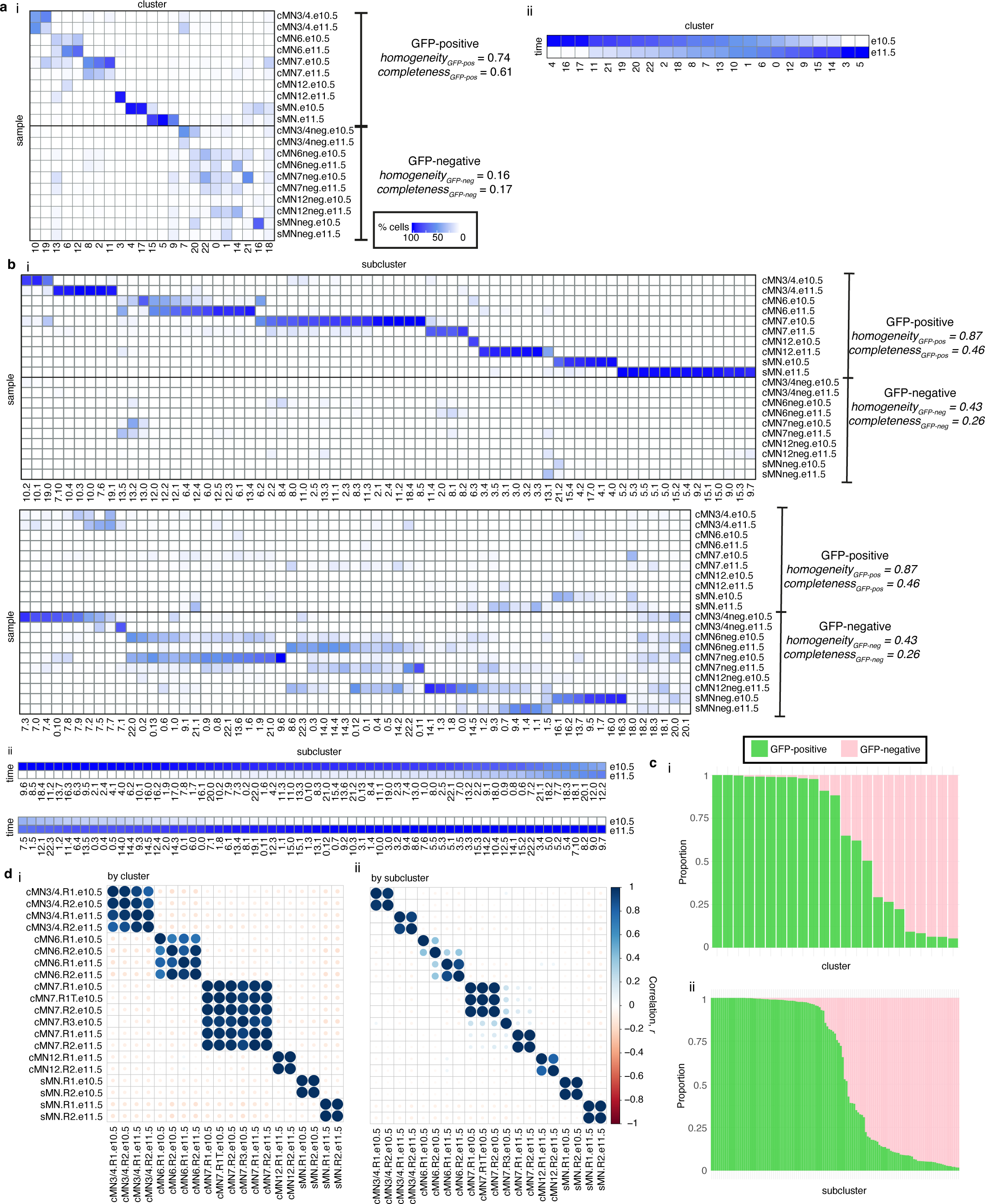
scATAC cluster purity across major clusters and subclusters. a. Heatmaps depicting purity of the 23 major scATAC clusters, stratified by i) sample and ii) embryonic age. cMN7 cells migrate past cMN6, are in close spatial proximity at these developmental ages, and are commonly co-dissected. Samples are GFP-positive unless otherwise marked (‘neg’). Clusters with higher membership from GFP-positive samples have higher purity than clusters with higher membership from GFP-negative samples. Most clusters feature cells from both e10.5 and e11.5 dissections, consistent with ongoing cell birth and proliferation. Homogeneity/completeness metrics calculated for GFP-positive versus GFP-negative samples are shown. b. Heatmaps depicting purity of the 132 scATAC subclusters, stratified by i) sample and ii) embryonic age. As observed with the major clusters in (a), subclusters with high GFP-positive membership have greater purity than high GFP-negative subclusters. In contrast to the major clusters, a greater proportion of subclusters have skewed temporal membership (e10.5 vs. e11.5), potentially reflecting transient cell states. c. Stacked barplots depicting proportion of GFP-positive and -negative cells within each i) cluster and ii) subcluster. Most clusters and subclusters are skewed towards pure (i.e., > 90%) GFP-positive or -negative membership. Here Cluster/subcluster IDs are not shown for ease of visualization. Detailed cluster annotations are available in **Supplementary Table 3**. d. Correlation matrix depicting pairwise correlations between all biological replicates among i) major clusters and ii) subclusters. Cluster/subcluster membership is highly correlated across biological replicates from different batches, particularly for subclusters.

**Extended Data Figure 5.**
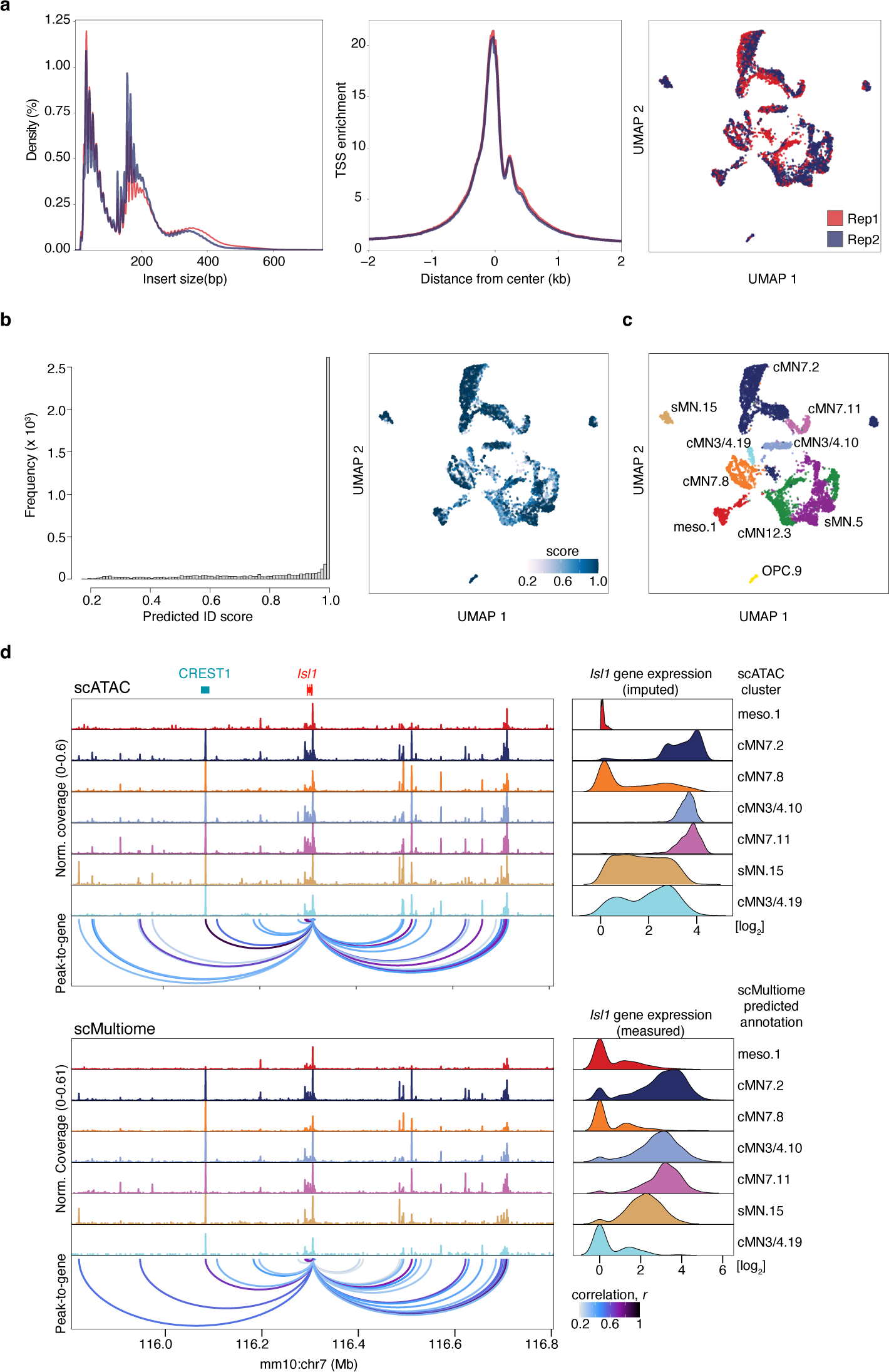
Single cell multiome reproducibility and quality control. a. Chromatin fragment length distribution (left), transcription start site (TSS) enrichment (middle), and joint UMAP embedding (right) comparing scMultiome biological replicates (red and blue). Replicates are highly concordant. b. Histogram (left) and UMAP embedding (right) depicting distribution of scMultiome prediction ID scores of annotations transferred from the scATAC reference set to the scMultiome query set using the *TransferData()* function in Seurat^128^. The distribution is heavily skewed towards higher scores. c. scMultiome annotations based on prediction IDs. Most predicted annotations correspond to *Isl1^MN^*:GFP-positive cell types, consistent with scMultiome dissection strategy. d. Direct comparison of peak-to-gene links from scATAC versus scMultiome for motor neuron master regulator *Isl1*. scATAC peak-to-gene links are generated from imputed gene expression values (“GeneIntegrationMatrix”) whereas scMultiome links are generated from direct gene expression measurements (“GeneExpressionMatrix”). Ground truth enhancer CREST1 is highly accessible in *Isl1*-positive clusters with strong peak-to-gene links across both modalities.

**Extended Data Figure 6.**
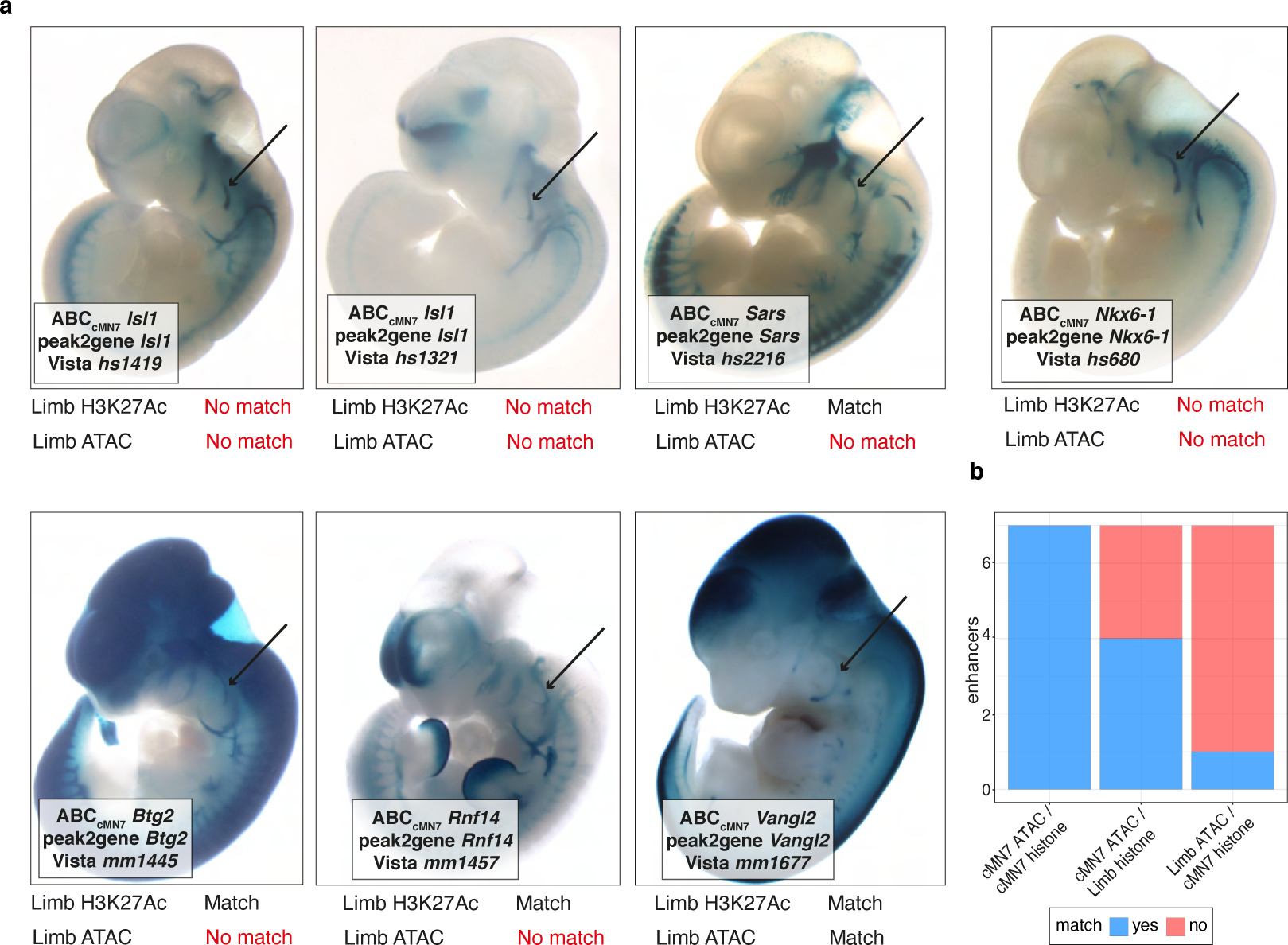
Toggling input data for Activity-by-Contact enhancer prediction. a. Whole mount *in vivo* enhancer reporter expression for the seven VISTA Enhancers that are annotated for cranial nerve (CN) expression, inspected for and have CN7 expression, and have positive Activity-by-Contact (ABC) enhancer predictions for CN7 at e11.5. Peak-to-gene predictions match ABC predictions in all cases (7/7). Replacing CN7 e11.5 H3K27Ac or ATAC data with these data from a distantly related cell type (mouse embryonic limb e11.5) results in either a matching or a non-matching cognate gene prediction. Substituting cMN7 e11.5 histone modification data with “Limb H3K27Ac” histone modification data alters predictions for 3 out of 7 enhancers. Substituting cMN7 scATAC data with “Limb ATAC” data alters predictions for 6 out of 7 enhancers. Neither substituted input correctly identifies the CREST1 enhancer (VISTA enhancer hs1419). Positive evidence of CN7 enhancement is depicted by arrows. b. Stacked barplot summarizing consequences of toggled input data.

**Extended Data Figure 7.**
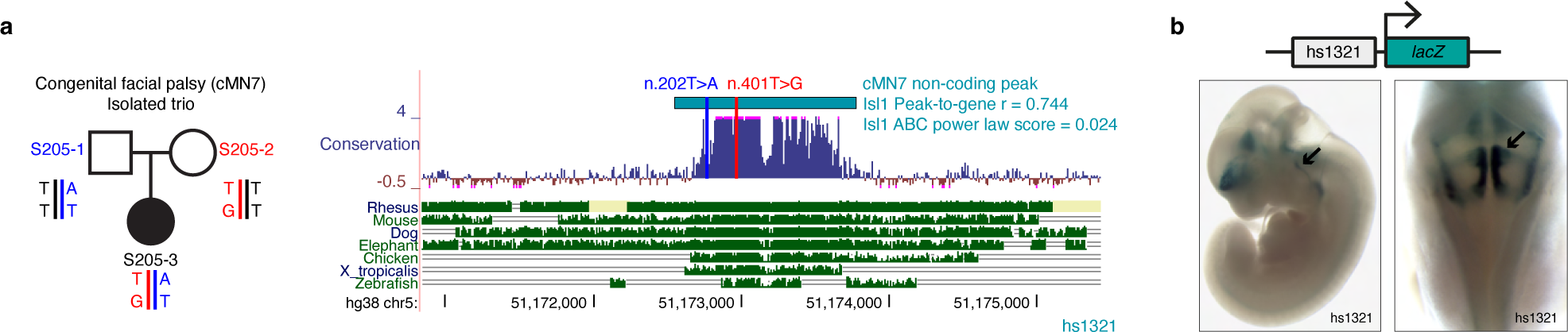
Compound heterozygous non-coding candidate variants in an *ISL1* enhancer. a. An affected trio with isolated congenital facial palsy, a CCDD affecting cMN7 (left), in which the affected offspring harbors compound heterozygous non-coding candidate SNVs (depicted by blue and red bars) affecting highly conserved nucleotides in enhancer hs2757 (right). The enhancer is predicted to regulate *Isl1* (peak-to-gene *r* = 0.744, ABC power law = 0.024). Variant coordinates are in NG_023040.1. b. *In vivo* reporter assay testing hs2757 enhancer activity. Enhancement is present in cranial nerve 7 (arrows), an *Isl1* positive cell type. Reporter expression views are shown as lateral (left) and dorsal through the 4^th^ ventricle (right).

**Extended Data Figure 8.**
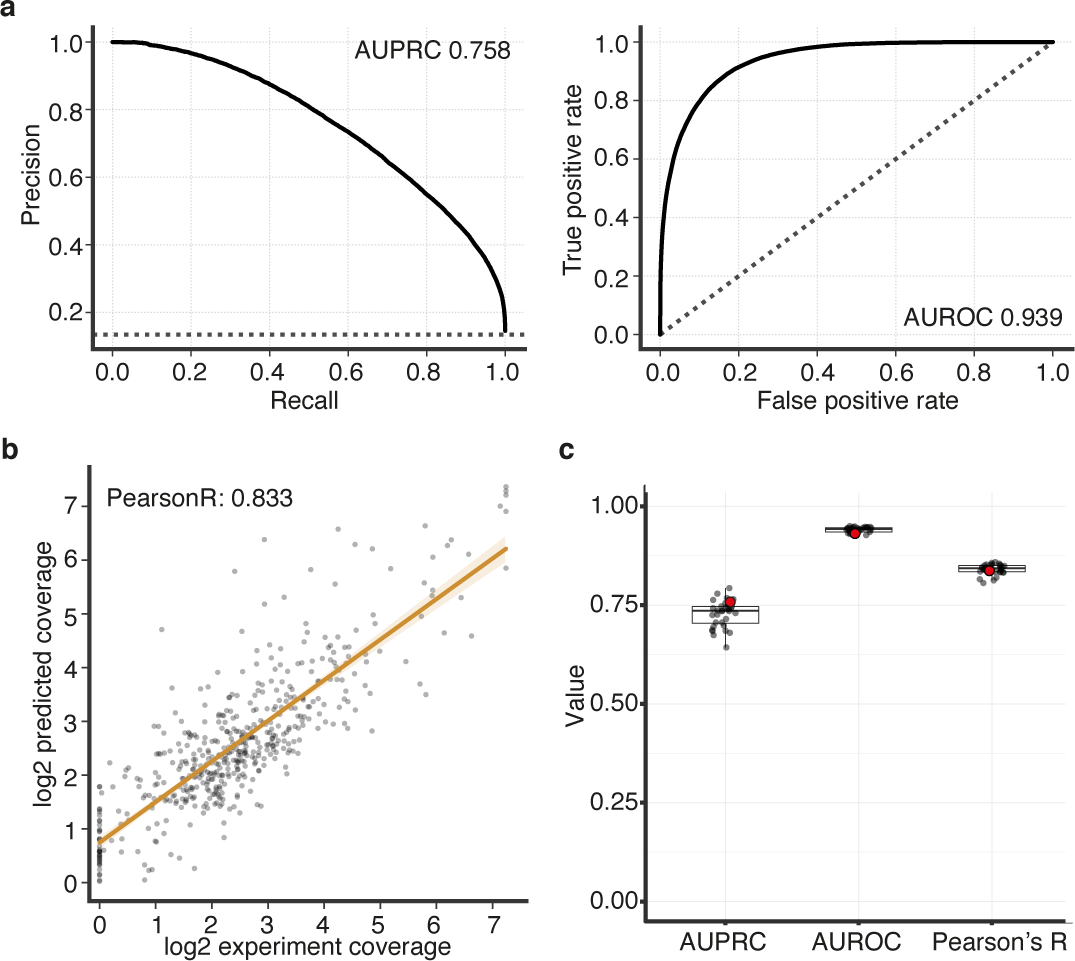
Quality metrics for *Basenji* convolutional neural network accessibility predictions. a. Precision-recall (PRC, left) and receiver-operating characteristic (ROC, right) curves measuring favorable performance (as measured by positive predictive value, sensitivity, true positive rate, and false positive rate) of *Basenji* accessibility predictions for cMN7 e10.5. AU denotes area under curve. Dotted lines represent the baseline classification rate. b. Scatterplot depicting *Basenji* accessibility predictions vs. true scATAC sequencing coverage for cMN7 e10.5. Each point represents a 128 bp test bin whose sequence was excluded from training. Measured and predicted coverage are positively correlated (Pearson’s R = 0.833). c. Boxplot summarizing area under PRC (AUPRC) and ROC (AUROC), and Pearson’s R for all samples and replicates. Quality metrics are consistent across samples. Data points depicted in (a) and (b) are highlighted in red. Centre line – median; box limits – upper and lower quartiles; whiskers – 1.5 x interquartile range.

**Extended Data Figure 9.**
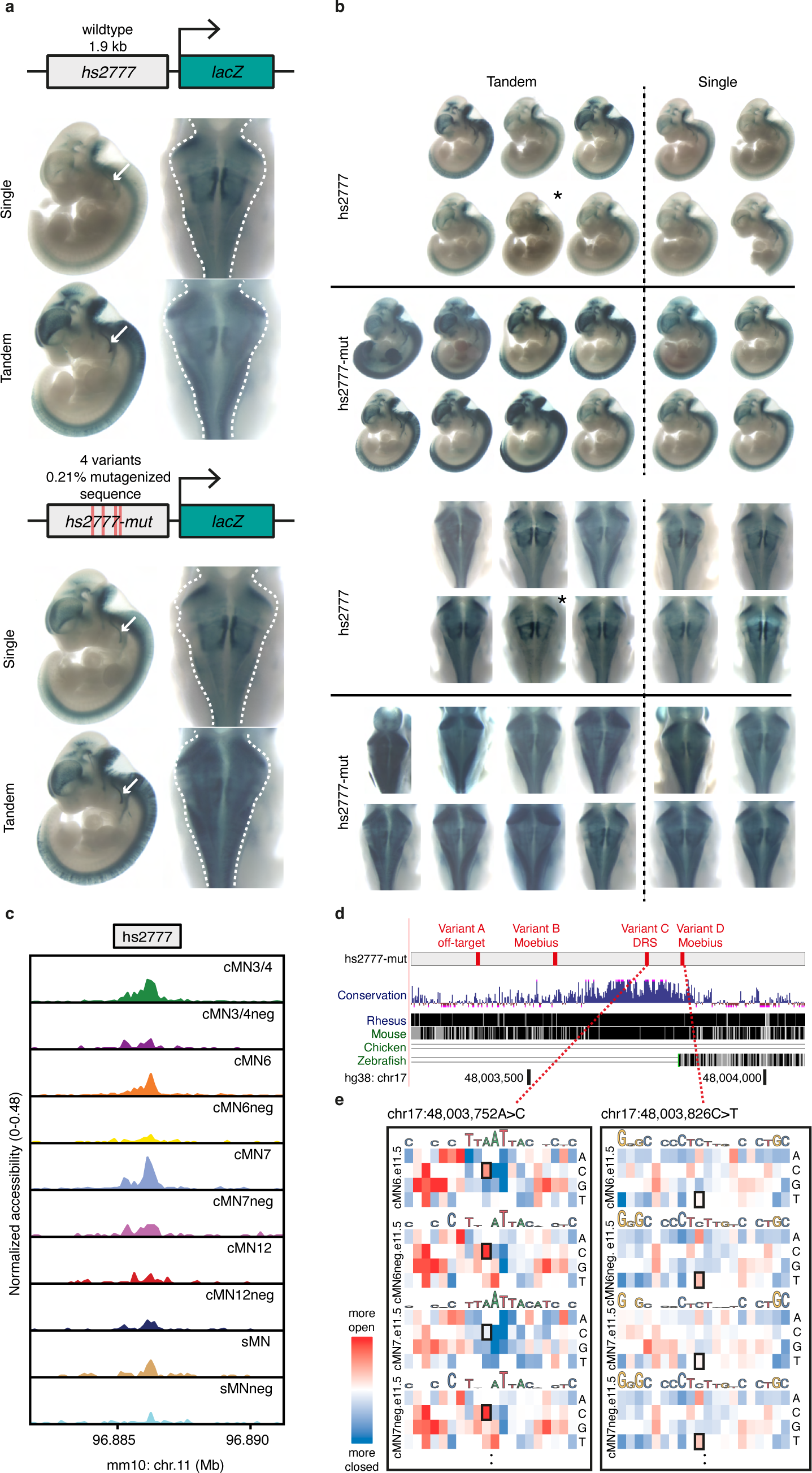
Cell type-aware candidate variants alter reporter expression *in vivo.* a. Representative whole mount *in vivo* enhancer reporter expression for (top) hs2777 wildtype and (bottom) hs2777-mut enhancer constructs. For each reporter insertion, dosage is labelled (“single”, “tandem”). Reporter expression views are shown as lateral (left) and dorsal through the 4^th^ ventricle (right). Cranial nerve 7 (white arrows) and surrounding hindbrain tissue (dashed lines) show visible gain of reporter expression. b. Additional replicates as in (a), matched by injection batch (top and bottom). hs2777-mut constructs reproducibly show increased reporter expression across midbrain, hindbrain, and neural tube. Random insertions are denoted by an asterisk. c. hs2777 chromatin accessibility profiles in the cranial motor neurons and surrounding cell types. The wildtype element is accessible across multiple cMNs and surrounding cells. d. UCSC screenshot depicting location of hs2777-mut variants: “Variant A” (chr17:48003393G>A, off-target), “Variant B” (chr17:48003557C>G, Moebius), “Variant C” (chr17:48003752A>C, DRS), and “Variant D” (chr17:48003826C>T, Moebius). hs2777-mut overlaps conserved non-coding sequence, particularly for Variants C and D. e. Neural net-trained *in silico* saturation mutagenesis predictions for all possible nucleotide changes in hs2777 for selected samples cMN6 e11.5, cMN6neg e11.5, cMN7 e11.5, and cMN7neg e11.5. Predicted loss-of-function nucleotide changes are colored in blue and gain-of-function in red. Specific nucleotide changes corresponding to *in vivo* Variants C and D are boxed. Samples marked with “neg” are GFP-negative cells surrounding the motor neurons of interest. All other samples are GFP-positive motor neurons. Variants C and D are predicted to increase accessibility in relevant samples consistent with their corresponding phenotypes; DRS alters cMN6 but not cMN7 development (Variant C), while MBS alters both (Variant D).

## METHODS

### Mouse husbandry, dissection, dissociation, FACS

We performed husbandry, dissection, dissociation, and fluorescence-activated cell sorting (FACS) as described previously^128^. Briefly, we crossed C57BL/6 (JAX # 000664) female mice with either 129S1/C57BL/6J *Isl^MN^:*GFP (JAX # 017952^35^) or *Hb9*:GFP (JAX # 005029^128^) male mice and separated them following one night of breeding. Pregnant females were sacrificed at 10.5 or 11.5 days post-conception and whole embryos were grossly dissected in chilled 1× PBS (ThermoFisher) then immediately placed in 1× B27 supplement (Gibco 17504044) in Hibernate E (Fisher NC0285514). Next, GFP-positive cranial motor neurons, GFP-positive spinal motor neurons, and GFP-negative surrounding cells were microdissected in pre-chilled HBSS (ThermoFisher) and placed in 1× B-27 supplement, 1× Glutamax (ThermoFisher 35050061), and 100 U/mL Penicillin-Streptomycin (PenStrep, ThermoFisher 15140122) in Hibernate E (medium 2). Microdissected tissues were dissociated using papain and ovomucoid solutions prepared from Papain Dissociation System (Worthington Biochemical LK003150). Tissues were resuspended in papain solution. Samples were then incubated at 37°C for 30 minutes and agitated every 10 minutes to ensure complete dissociation. Following incubation, samples were spun down at 300 rcf for 5 minutes, the supernatant was removed, and dissociated tissues were resuspended in 500 uL of ovomucoid solution (plus or minus 100 μL depending on quantity of tissue). Tissues were again spun down at 300 rcf for 5 minutes and resuspended in 500 μL of medium 2 (plus or minus 100 μL depending on quantity of tissue) and transferred to a 5mL polystyrene round bottom tube on ice. Live GFP-positive singlets were separated from GFP-negative cells (GFP-negative limb buds from embryos used as negative control to set gates) using an ARIA-561 FACS machine at the Immunology Research Core at Harvard Medical School (for ATAC-seq samples), and an BD FACS Aria II at the Jimmy Fund Core at the Dana-Farber Cancer Institute (for bulk and single cell RNA-seq samples). GFP-positive cells were collected either into 200 uL of media containing 1× Glutamax, 100 U/mL PenStrep, and 2% 2-Mercaptoethanol (Gibco 21985023) in Neurobasal-A Medium (ThermoFisher 10888022) for ATAC-seq, or into 96 well fully-skirted Eppendorf plates containing a starting volume of 5 ul/well of Hibernate E for single cell RNAseq, or directly into 1.5 ml tubes containing Qiagen RNeasy Lysis buffer/Buffer RLT (Qiagen 79216) for the bulk RNAseq. Embryos were not selected based on sex. Embryos were excluded if they did not match expected developmental stage as estimated from morphological features.

### Single cell ATAC-seq: Nuclei Isolation, tagmentation, and sequencing

We performed fluorescence-assisted microdissection to collect samples cMN3/4, cMN7, and sMN from *Isl1^MN^*:GFP mice and likewise to collect samples of cMN6, cMN12, and sMN from *Hb9*:GFP mice, each at both e10.5 and e11.5. We performed FACS-purification as described above to collect GFP-positive motor neurons, as well as GFP-negative cells surrounding the motor neurons to better distinguish between motor neuron versus non motor neuron regulatory elements (for a total of 20 sample types, 9 with biological replicates and 2 with technical replicates for 32 samples in all). Nuclei were isolated in accordance with Low Cell Input Nuclei Isolation guidelines provided by ‘Demonstrated Protocol – Nuclei Isolation for Single Cell ATAC Sequencing Rev A’ from 10x Genomics. Cell suspensions were spun down at 300 rcf for 5 min at 4°C in a fixed angle centrifuge, the supernatant was removed, and the pellet was resuspended in 50 uL of 0.04% BSA in PBS. The cell solution was then transferred to 0.2 mL tube and centrifuged at 300 rcf for 5 minutes at 4 °C in a swinging bucket centrifuge. Without contacting the bottom of the tube, 45 uL of supernatant was removed, and the cell pellet was resuspended in 45 uL of chilled Lysis buffer (10 mM Tris-HCl (pH 7.4), 10 mM NaCl, 3 mM MgCl_2_, 0.1% Tween-20, 0.1% Nonidet P40 Substitute, 0.01% Digitonin, 1% BSA, in nuclease-free water). Nuclei suspensions were incubated on ice for 3 minutes and 50 uL of wash buffer (10 mM Tris-HCl (pH 7.4), 10 mM NaCl, 3 mM MgCl_2,_ 1% BSA, 0.1% Tween-20, in nuclease free water) was added to the suspensions without mixing. Nuclei suspensions were then spun down in a swinging bucket centrifuge at 500 rcf for 5 minutes at 4 °C, 95 uL of supernatant was removed, and 45 uL of nuclei buffer was added. Samples were again spun down in a swinging bucket centrifuge at 500 rcf for 5 minutes at 4 °C, all supernatant was removed without contacting the bottom of the tube, and nuclei were resuspended in 7 uL of nuclei buffer. 2 uL of this final nuclei suspension was added to 3 uL of nuclease-free water, and 5 uL of trypan blue, and cell viability was inspected using the Countess II FL Automated Cell Counter (Thermo Fisher Scientific AMQAF1000). We performed scATAC transposition, droplet formation, and library construction as described in protocol CG000168 using v1 reagents (10x Genomics). scATAC libraries were sequenced on the Illumina NextSeq 500 system using standard Illumina chemistry. Paired inserts were minimum 2 x 34 bp in length excluding indices, and libraries were distributed to achieve an estimated coverage of ≥ 25,000 read pairs per cell in accordance with 10x Genomics guidelines (actual mean coverage was 48,772 reads per cell). Samples failing quality control were excluded (e.g., failed TapeStation output).

### scATAC preprocessing, peak calling, dimensionality reduction, and cluster analysis

We performed a modified workflow based on Cusanovich *et al.*^129^. Briefly, we generated fastq files from bcl using cellranger *mkfastq*. We initially included all single cell ATAC barcodes perfectly matching an allowlist provided by 10x Genomics. We also included fixed barcodes if they had a maximum Hamming distance of 1 and if they were present in the top 2% of barcode counts. As a final check, we manually inspected the distribution of fixed barcodes in reduced dimension space to ensure a roughly even distribution across all cells. We aligned individual samples to the mm10 reference genome using Bowtie2^129^, generated sample level .bam files, filtered reads with MAPQ < 10, and performed PCR deduplication. We established heuristic coverage per cell thresholds for each sample separately. To generate cell counts, we performed hard filtering based on log10[nfrags/barcode] for each sample separately.

We performed LSI-based clustering to generate sample-level clades as described previously^130^. In order to enrich peak representation from rare neuronal populations, we manually assigned between 3-7 clades to each sample and then performed peak calling on each clade using MACS2^130^. We first performed cell QC based on heuristic filters (low FRiP and accessible peaks-per-cell outliers), then peak QC (filtering peaks in a low proportion of remaining cells per clade). All post-QC cells and peaks were then combined to generate a master peak-by-cell callset. Samples failing any stage of QC were excluded (e.g., inadequate read coverage).

We performed LSI-based dimensionality reduction (log-scaled TF-IDF transformation followed by singular value decomposition) on our binarized peak-by-cell matrix as based on previously described methods^130^. We used *umap()* (https://github.com/lmcinnes/umap) to further reduce the dimensionality of our data to 3-dimensional UMAP coordinates. We then performed cluster analysis using Seurat’s SNN-graph approach. Once the major clusters were defined, we repeated our dimensionality reduction and cluster analysis on each major cluster to generate subclusters.

### Cluster homogeneity, completeness, and purity

In order to formalize the agreement between our dissection/FACS labels (“class”) and our cluster/subcluster labels (“cluster”), we calculated homogeneity *h*, completeness *c*, and Vmeasure *V_β_*, using the *sabre* package^131^:

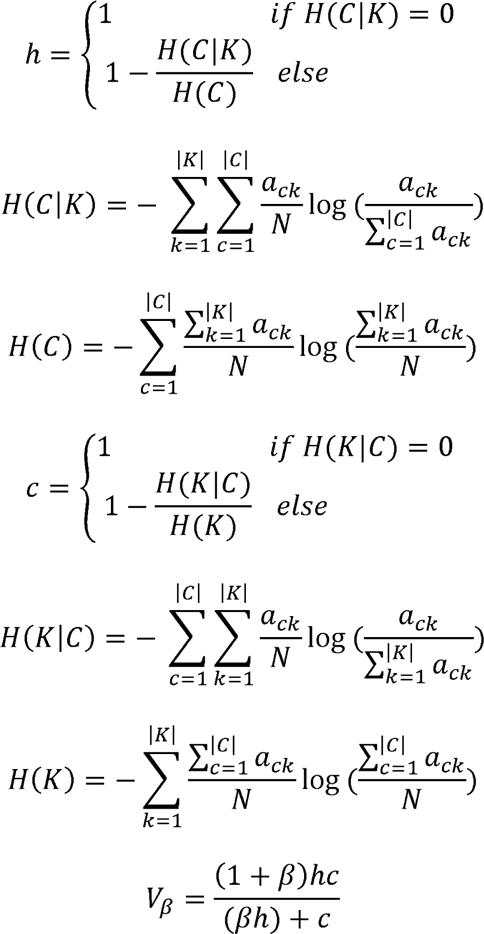

Where *C* is the set of dissection/FACS class labels; *K* is the set of clusters or subclusters; *a_ck_* is the number of single cells belonging to class *c* and cluster or subcluster *k*; *N* is the total number of single cells; and *β* is the ratio of weights attributed to *c* and *h* (*V_β_* is the weighted harmonic mean of *c* and *h*). As *β* becomes very large or very small, *V_β_* approaches *c* and *h*, respectively. Here we set *β* to 1.

We also generated a per-cluster purity metric, *p* to quantify the maximum cellular representation of each cluster/subcluster:

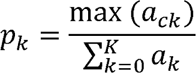

Homogeneity, completeness, and Vmeasure calculations across varying conditions of *C* and *K* are summarized in **Supplementary Table 4**.

### Motif Enrichment and aggregated footprinting analysis

We used the mouse motifs from the cisBP database from the chromVARmotifs database to compute cluster and sample specific motif footprinting and enrichments (mouse_pwms_v2). For each motif, we identified all sites in peaks where a motif was present. Clusters 3, 4, 5, and 9 were excluded from footprint analysis. We next identified differentially accessible peaks for each group of interest using ArchR’s *getMarkerFeatures()* function, normalizing for differences across groups with transcriptional start site (TSS) Enrichment and log10(nFrags). We selected peaks for each group that met an FDR threshold of below 0.01 and a LogF2C of >=1. Aggregated footprint plots were generated for select motifs using *plotFootprints()*, by first normalizing the Tn5 bias by subtracting it from the footprinting signal. For site-specific footprints, we used TOBIAS to generate Tn5-bias corrected bigwigs and footprint scores across the genome for each cell type^131^. For bias estimation and correction we excluded ENCODE denylist regions from *mm10-blacklist.v2.bed* (https://github.com/Boyle-Lab/).

### *In vivo lacZ* enhancer validation

We selected 25 putative wildtype enhancers for downstream experimental validation based on the following criteria. First, we selected elements with significant cell type specificity scores^51^. Next, we excluded any elements that did not lift over to the human genome (hg19). We then identified elements with evidence of H3K27Ac marks in the ENCODE portal^131^ and no existing experimental data in the VISTA enhancer browser^132^ (freeze September 2019). Finally, we performed manual curation in order to select for elements with high conservation, against elements in repetitive regions, and ensured representation of elements from cMNs 3, 4, 6, 7, 12, and sMNs.

We performed *in vivo* enhancer testing using the enSERT transgenesis method described by Osterwalder *et al.*^133^. Briefly, the orthologous human sequence each candidate enhancer was cloned into a pCR4-Shh::lacZ-H11 vector (Addgene plasmid # 139098) containing the mouse *Shh* minimal promoter, *lacZ* reporter gene, and H11 safe harbor locus homology arms. The cloned construct, Cas9 protein, and H11-sgRNAs were delivered via mouse embryonic pronuclear injection (mouse FVB/NJ JAX #001800) and transferred to female hosts. Embryos were collected at e11.5, stained with X-gal, and evaluated for reporter activity.

For candidate variant testing, we generated enhancer clones bearing the human reference or variant allele as described above. In the case of compound heterozygous variants, we cloned both variants into the same construct in *cis.* In the case of full enhancer deletion candidates, we cloned only the wildtype enhancer.

### Bulk ATAC-seq

We performed bulk ATAC-seq as described previously^127^ for FACS-purified cells from six anatomic/temporal regions: *Isl^MN^*:GFP-positive cMN3 at e10.5 and e11.5, cMN7 at e10.5, sMN e10.5 and e11.5, and *Isl^MN^*:GFP-negative hindbrain at e11.5. We processed the bulk ATAC sequencing data by running the .fastq files through the Encode ATAC-seq pipeline (https://github.com/ENCODE-DCC/atac-seq-pipeline) using default parameters. To analyze peaks for each bulk sample, we used Irreproducible Discovery Rate (IDR) optimal peaks, generated between pseudoreplicates or biological replicates when appropriate. After generating peaksets for each bulk sample, we created a bulk master peakset by concatenating all the individual peaksets and merging with bedtools *merge*. We further generated bulk peaksets specific to each sample using bedtools *subtract*, allowing for ≤ 50% overlap between peaks.

### Single Cell RNA-seq

Husbandry and collection strategy was identical to the scATAC strategy described above, except that we combined GFP-positive and -negative cells from the same dissections. We performed single cell RNA-seq for FACS-purified eGFP-positive motor neurons from 6 anatomic/temporal regions: cMN3+4 and cMN7 from *Isl1^MN^*:GFP mice and cMN6 from *Hb9*:GFP mice, all at both e10.5 and e11.5 (for total of 10 samples). In most samples we spiked in 10% surrounding eGFP-negative hindbrain cells as an internal control for comparison to non-motor neurons. Samples were submitted to the Klarman Cell Observatory/Regev Lab at the Broad Institute of MIT and Harvard for processing on a 10X Genomics Chromium platform. The 10X Genomics Chromium Single Cell 3’ Reagent Kit (using v2 single index chemistry, CG00052) was used for mRNA capture and library preparation. Samples were multiplexed for a read-depth goal of 50,000 reads/cell (actual mean coverage was 94,829 reads/cell). Sequencing was performed on a HiSeq 4000 by Broad Genomic Services using standard Illumina chemistry. The data was then aligned in the Engle lab using Cell Ranger v2.1.1 against the ENSEMBL Mus musculus genomic reference build GRCm38.87 (modified to include eGFP and tdTomato sequences). Quality control was performed in Seurat to remove doublets and low-read cells. Analysis was done in Seurat where samples were integrated with Canonical Correlation Analysis (CCA)^134^. Motor neurons were identified from *eGFP, Isl1* and expression of other motor neuron markers (eGFP was regressed out to avoid affecting clusters),

### Bulk RNA-seq

We performed bulk RNA-seq for FACS-purified eGFP+ cells from 7 anatomic/temporal regions: cMN3, cMN4, cMN6, cMN7 at each corresponding brainstem level, at both e10.5 and e11.5 (except for cMN6 that was only collected at e11.5 due to cell number limitations at e10.5; with two biological replicates from all times/regions and 1 additional technical replicate of cMN6, for a total of 15 samples). Samples from multiple litters were merged to reach a threshold for appropriate cell number and sent to Rutgers RUCDR for library preparation and sequencing. For the e11.5 samples, 200 ng/sample of RNA was isolated with Oligo-dT beads, enriching for mRNA. Depletion of beta globin mRNA and ribosomal RNA was performed. For the e10.5 samples and the e11.5 cMN6 samples, due to the lower total RNA from fewer starting cells in these nuclei at these ages, whole-transcriptome Nugen Amplification was performed. Samples were sequenced with a 100 bp paired-end strategy to sequence full-length transcripts on an Illumina HiSeq2500 for an approximate read-depth of 60 million paired-end reads/sample. This generated R1 and R2 reads for each of 2 lanes of data/sample that were subsequently concatenated. STAR (Spliced Transcripts Alignment to a Reference)^134^, a splice-aware tool, was used to align reads to ENSEMBL Mus musculus genomic reference build GRCm38.87, and RSEM (RNA-Seq by Expectation Maximization)^135^ was used to generate the count files. We then used DESeq2^136^ to make comparisons.

### Generating peak-to-gene links

For our original RNA inputs for peak-to-gene links, we performed scRNA-seq on cMN3+4, cMN6, and cMN7 dissections (GFP-positive and -negative) at e10.5 and e11.5. Our husbandry and collection strategy was identical to the scATAC strategy described above, except that we combined GFP-positive and -negative cells from the same dissections. We performed scRNA seq as described in protocol CG000168 using v2 single index chemistry and sequenced on the Illumina HiSeq 4000. To benchmark our scRNAseq results, we also performed bulk RNAseq on cMN3, cMN6, and cMN7.

We integrated multiple scRNA-seq datasets from GFP-positive and -negative cells from cMN3/4, 6, and 7 dissections at e10.5 and e11.5 into a single Seurat object using Seurat’s integration framework^76,135^. We excluded cells with more than 5% of reads aligning to the mitochondrial genome. After examining the distribution of the number of unique features and number of unique reads per cell for each sample, we manually filtered cells with low feature counts. Finally, we normalized each sample using the *NormalizeData()* function, identified the top 10,000 variable features per sample, and scaled each sample using the *ScaleData()* function.

Next, we excluded scATAC clusters (clusters 3, 4, 5, and 9) with high proportions of GFP-positive sMN and cMN12 dissected cells, as those samples are not represented in our scRNA dataset. We then performed unconstrained scATAC-RNA integration on all remaining cells using *addGeneIntegrationMatrix()* in ArchR^135^.

We then evaluated the projected gene expression values from our scATAC-RNA integration for three high-confidence scATAC clusters (cMN3/4.10, cMN6.6, and cMN7.2). We selected these clusters due to unambiguous sample membership based on microdissection origin (purity), FACS labels (corresponding to cMN7, cMN6, and cMN3/4, respectively), and known marker locus accessibility/expression. We compared imputed gene expression from these clusters to corresponding bulk RNAseq samples that were independently dissected and FACS purified. Specifically, we performed differential expression analysis on bulk RNAseq data (DEseq v1.34.0^136^) and on imputed gene expression on scATACseq data (using *getMarkerFeatures()* function in ArchR). We fit a linear model of the log_2_[fold-change] expression for all combinations of bulk samples and single cell clusters, and confirmed a significant positive correlation between projected gene expression for marker genes in each cluster against its corresponding bulk counterpart.

We calculated peak-to-gene correlations using ArchR’s *addPeak2GeneLinks()* function, with reducedDims = “IterativeLSI_ArchR’’. We included all high confidence links (FDR < 0.0001) with a minimum correlation coefficient of ≥ 0.1, within +/- 500 kb of a given gene, which we reasoned would include the vast majority of putative enhancers^76,137^, including those active in only a subset of cells.

We then benchmarked this cMN peak-to-gene set against two alternative scATAC-RNA integrations using subsetted scRNAseq data from the Mouse Organogenesis Cell Atlas (MOCA)^137^. First we created a neuronal dataset set by integrating our oversampled cMN scATAC profiles with more uniformly sampled sci-RNA neuronal clusters from MOCA (annotated as “Cholinergic Neurons”, “Excitatory Neurons”, “Inhibitory Neurons”, “Neural Progenitor Cells”, “Postmitotic Premature Neurons”, “Primitive Erythroid Lineage”, and “Stromal Cells”). We removed any cells that were not collected at e10.5 and e11.5 to age-match our scATAC set. We also performed an scATAC-RNA integration using a more distantly related cell type with minimal sampling overlap, (sci-RNA MOCA Cluster 34 annotated as “Cardiac Muscle Lineage”) and included non-age-matched cells for this integration. We then generated peak-to-gene links as described above and quantified the total number of links across different RNA integrations.

To quantify and compare the distribution of peak-to-gene links across different genes, we tabulated significant peak-to-gene links (r > 0.1 and FDR < 10^-4^) +/- 50 kb of each gene’s TSS. In the case of peaks connected to multiple genes, we selected the link with the lowest FDR value. Next, we generated modified Domain of Regulatory Chromatin (DORC) scores first described by Ma *et al.*^138^ by normalizing all reads in our peak-by-cell matrix by unique fragment count. We then summed these normalized values for all peak-to-gene connections within +/- 500 kb of each gene TSS for every cell.

### Single cell Multiome (scMultiome)

We performed timed matings, microdissections, dissociation, and FACS to collect GFP-positive cMN3/4, cMN7, cMN12, and sMN cells at e11.5 as described above. Instead of generating separate reactions for each cell type, we pooled these cells prior to dissociation, selected GFP-positive cells via FACS, and performed Low Cell Input Nuclei Isolation (10x Genomics CG000365) and Single Cell Multiome ATAC + Gene Expression assay (10x Genomics CG000338) on a total of two pooled replicates. We performed sequencing on a NextSeq 500 for Multiome ATAC and Gene Expression libraries separately, using a custom sequencing recipe for ATAC provided by Illumina. We performed QC, dimensionality reduction, and generated peak-to-gene links as described above using functionality in Signac and ArchR^70,139^. In order to facilitate direct comparison across modalities, we calculated scMultiome fragment depth against our high confidence scATAC peakset. We calculated multimodal weights for each cell using a weighted nearest neighbour approach^140^ and performed *ab initio* graph-based clustering on our scMultiome cell set. In order to annotate these clusters, we generated cell-cell anchors by defining scMultiome clusters as the query set and our well-annotated scATAC clusters as the reference set. Because each multiome cluster was typically dominated by a single predicted scATAC cluster, we annotated each multiome cluster based on its maximum predicted scATAC membership.

### Single cell CUT&Tag

We collected cranial motor neurons (GFP-positive cMN3+cMN4 e11.5, cMN6 e11.5, cMN7 e10.5, and cMN7 e11.5) as described above and performed a modified scCUT&Tag protocol74,125. Briefly, we collected GFP-positive cells directly into fresh antibody buffer (2011mM HEPES pH117.5, 15011mM NaCl, 0.511mM spermidine, 1× protease inhibitor (Sigma 11873580001), 2 mM EDTA, 0.05% digitonin, 0.01 % NP-40, 1× protease inhibitors and 2% filtered BSA). We centrifuged samples at 450 rcf for 5 minutes, washed in 200 uL antibody buffer, centrifuged at 600 rcf for 3 minutes, resuspended in 1:50 H3K27Ac primary antibody (monoclonal Rabbit anti-mouse, Abcam ab177178), and incubated overnight at 4°C with gentle rotation. Nuclei were centrifuged at 600 rcf for 3 minutes, washed in 200 uL Dig-Wash-BSA buffer (2011mM HEPES pH117.5, 15011mM NaCl, 0.511mM spermidine, 1× protease inhibitor, 0.05% digitonin, 0.01 % NP-40, 1× protease inhibitor and 2% filtered BSA), centrifuged at 600 rcf for 3 minutes, resuspended in 1:50 IgG secondary antibody (guinea pig anti-rabbit Novus Biologicals, NBP1-72763), and incubated 1 hour at room temperature with gentle rotation. Nuclei were then centrifuged at 600 rcf for 3 minutes, washed 3x in Dig300-Wash-BSA (2011mM HEPES pH117.5, 300 mM NaCl, 0.511mM spermidine, 1× protease inhibitor, 0.05% digitonin, 0.01% NP-40, 1× protease inhibitors and 2% filtered BSA), resuspended in 1:20 pAG-Tn5 (EpiCypher 15-1017), and incubated 1 hour at room temperature with gentle rotation. Nuclei were centrifuged at 450 rcf for 3 minutes, washed 3x in Dig300-Wash-BSA, resuspended in 200 uL tagmentation buffer (2011mM HEPES pH117.5, 30011mM NaCl, 0.511mM spermidine, 1× protease inhibitor, 0.05% digitonin, 0.01 % NP-40, 1× protease inhibitor, 2% filtered BSA, and 10 mM MgCl2), incubated 1 hour at 37°C with agitation every 15 minutes. Tagmentation was halted with Stop buffer (2011mM HEPES pH117.5, 300 mM NaCl, 0.511mM spermidine, 1× protease inhibitor, 0.05% digitonin, 0.01% NP-40, 1× protease inhibitors, 2% filtered BSA, and 25 mM EDTA), centrifuged at 450 rcf for 3 minutes, washed in diluted nuclei buffer (1× ATAC Nuclei Buffer (10x Genomics, PN-2000207) and 2% filtered BSA), centrifuged at 450 rcf for 3 minutes, and resuspended in diluted nuclei buffer. Intact nuclei were stained with DAPI and were visualized and counted under fluorescent microscopy. 70 uL of ATAC master mix (8 μL tagmented nuclei, 7 μL ATAC Buffer B (10x Genomics, PN-2000193), 56.5 μL Barcoding Reagent B (10x Genomics, PN-2000194), 1.5 μL Reducing Agent B (10x Genomics, PN-2000087), 2 μL Barcoding Enzyme (10x Genomics, PN-2000139) was loaded for GEM generation according to the 10x Genomics scATAC v1.1 protocol. Nuclei were diluted if necessary (up to a maximum of 25,000 total nuclei per reaction). Subsequent GEM generation and cleanup steps were performed according to the 10x Genomics scATAC v1.1 protocol. Library prep was also performed using the standard protocol, except that total PCR cycles were increased to 16. All centrifugation steps were performed using a swing-bucket rotor.

### Activity-by-contact (ABC) enhancer predictions

We generated enhancer predictions for four cell types, GFP-positive cMN3+4 e11.5, cMN6 e11.5, cMN7 e10.5, and cMN7 at e11.5, adapting the Activity-By-Contact (ABC) model v0.2 described previously^139,140^. We defined potential enhancer regions by merging scATAC peaksets for each sample. We provided sample-specific H3K27Ac read counts from scCUT&Tag experiments described above. We also provided imputed RNA expression tables for each cell type from the scATAC-scRNA integration described above. We estimated contact frequencies based on the ABC power law function. We evaluated our enhancer predictions against 67 VISTA enhancers classified as positive for “cranial nerve”, of which 12 had ABC enhancer predictions. Importantly, our ABC predictions also correctly identify the peak and cognate gene for the CREST1 enhancer (VISTA enhancer hs1419), for which both the enhancer locus and cognate gene are known^140^.

### Participant whole genome sequencing, reprocessing, SNV/indel calling and quality control

Research participants were enrolled into the long-term genetic study of CCDDs at Boston Children’s Hospital (BCH; clinicaltrials.gov identifier NCT03059420). The Institutional Review Board at BCH approved the study. Informed consent was obtained from each participant or legal guardian. Individual-level data was de-identified and studies were performed in compliance with US 45.CFR.46 and the Declaration of Helsinki. WGS was performed at Baylor Human Genome Sequencing Center through the Gabriella Miller Kids First Pediatric Research Program (dbGaP Study Accession: phs001247). Joint variant calling for all samples was performed at the Broad Institute. We uploaded raw 30X coverage PCR-free WGS data to the Broad Institute’s secure Google Cloud server and reprocessed these data through the Broad Institute’s production pipeline. We realigned raw read data to the GRCh38 human reference sequence using BWA-MEM and reprocessed using the Broad’s Picard Toolkit. We then performed variant calling on the resultant BAM files using the Genome Analysis Toolkit (GATK 4.0 HaplotypeCaller). In the final step of variant calling, we jointly genotyped each site in the genome alongside a collection of over 20,000 reference genomes assembled by the Broad Institute. Joint variant calling provides two crucial advantages over individual or batched genotyping^141^. First, it dramatically improves variant calling accuracy due to i) clearer distinction between homozygous sites versus missing data; ii) greater sensitivity to detect rare variants, and iii) greater specificity against spurious variants. Second, joint calling by its design generates a well-calibrated estimate of allele frequency within our cohort against the large gnomAD database. Assuming that the allele frequency of a *bona fide* Mendelian disease-causing variant is lower than its disease prevalence, this information allows us to exclude variants with implausibly high allele frequencies^141,142^. Finally, we performed variant filtering using GATK’s Variant Quality Score Recalibrator and applied custom hard filters as required.

We performed rigorous QC at multiple stages of variant calling, performed filtering based on standard sequencing quality metrics (e.g., uniformity of coverage, transition/transversion ratio, indel length profiles, etc.), and compared them to our internal database of reference genomes. We used heterozygosity of common variants on chrX and coverage of sites on chrY to confirm reported gender and to identify sex chromosome aneuploidy. We also extracted variant calls from 12,000 well-covered variant sites and used these variants for principal component analysis together with a large reference panel to infer the geographical ancestry of samples, to infer pairwise relatedness of the samples, to identify unexpected duplicates, and to determine cryptic relatedness and unexpected patterns of relatedness within reported families. The data/analyses presented in the current publication have been deposited in and are available from the dbGaP database under dbGaP accession phs001247.v1.p1. Adult participants and guardians of children provided written informed consent for participation. No participant compensation was provided.

### Structural Variants

We generated an SV callset using the ensemble GATK-SV pipeline as described previously (https://github.com/broadinstitute/gatk-sv)^142–146^. Briefly, we performed joint genotyping and harmonized SV calls from multiple detection tools (Manta, Wham, MELT, GATK-gCNV, and cn.MOPS^143–147^), as well as manual read inspection using IGV^148^, and estimated SV allele frequencies against gnomAD SV v2.1. We first excluded any SVs with cohort AF ≥ 0.005, irrespective of coding or non-coding status. When evaluating for *de novo* and inherited SV candidates, we restricted our callset to 45 and 49 curated pedigrees, respectively. One SV (deletion chr22:27493955-27497536) was identified through manual curation. These SVs were subsequently used for downstream analysis incorporating pedigree non-coding element information.

We also performed a separate bespoke analysis for genome-wide transposon insertions (L1, Alu, and SVA) profiling on the GMKF WGS dataset using xTea^149^. Raw transposon insertions with different features and confidence levels were annotated and processed to generate both rare and *de novo* insertion lists for further variant interpretation. Beyond basic feature annotations (transposon family, breakpoint, and gene annotations), all insertions were annotated with 1) population allele frequencies (AFs) derived from the 1000 genomes project, gnomAD SV, euL1db, and other polymorphic insertion collections from the literature^81,150–152^; 2) overlapping repeats annotated by RepeatMasker and homopolymers; 3) other gene annotations such as pLI score, OMIM disease-causing genes, and potential CCDD-related genes. For putative pathogenic rare insertions, we first applied population AF threshold of 0.01 to remove common polymorphic insertions. We then filtered nested insertions–where a putative insertion landed in an existing insertion from the same transposon family–as they are error-prone in short read sequencing platforms. Finally, we filtered for all high confidence annotations (“two_side_tprt_both” and “two_side_tprt”) in affected samples for downstream genetic analysis. For *de novo* insertions, raw calls of transposon insertions were examined and only those present in the affected proband but fully absent in both parents (i.e., without a single supporting read) were retained. Trio families with any member bearing abnormal high number of transposon calls were filtered, as these outlier samples carried excessive noisy signals (clipped and discordant reads) and consequently false positive calls could affect *de novo* insertion calling. We then removed insertions that have been reported in populational datasets and known polymorphic insertion collections in the literature. We also filtered out error-prone nested insertions. Finally, high-confidence insertions (feature = “two_side_tprt_both”) in affected participants were reported as the *de novo* insertions for further genetic interpretation (**Supplementary Table 15**).

### Applying cell-type aware filters for human non-coding mutations

Our original WGS callset contained 49,824,956 variant calls for 899 individuals across 270 distinct families with CCDDs. We loaded these unfiltered variant calls in .vcf format into Hail (https://github.com/hail-is/hail) as a MatrixTable. Multi-allelic variants were split so that all variants are represented in a bi-allelic format. In splitting multi-allelic variants, spanning deletions were not kept. This resulted in 54,804,014 bi-allelic variants. These variants were annotated with TOPMed allele frequencies, gnomAD genomes allele frequencies and allele counts, GERP scores and ClinVar variant pathogenicity labels. Using native and custom Hail functions, we generated scripts to filter the MatrixTable’s variant calls based on custom specifications for variant annotations, variant locus, and call quality filters.

We set the following hard filters for all searches:

gnomAD AF^152^ (< 1 x 10^-3^ for dominant/de novo; < 1 x 10^-2^ for recessive)
TopMED AF^153^ (< 1 x 10^-3^ for dominant/de novo; < 1 x 10^-2^ for recessive)
GERP^154^ > 2
Only return variants that pass all quality filters in the VCF
Genotype Quality: > 20
Allele Balance: > 0.15 (heterozygous calls)

To generate a list of cell type specific genomic regions of interest for each disease group, we used data from single cell ATAC-seq experiments performed on mouse cranial motor neurons at e10.5 and e11.5. From here we implicitly assume that: i) we have correctly mapped each disease-relevant cell type (at the appropriate timepoint) to its appropriate cognate phenotype; ii) biologically active cREs are accessible; and iii) patterns of chromatin accessibility are correlated across species^148^. Peaks called on each cMN sample were lifted over from mm10 to hg38, and the converted intervals were concatenated into a single file and overlapping peaks were combined using bedtools *merge*. For disease types with > 1 cMN of interest, the master list of intervals for each cranial nerve were again merged using bedtools *merge* to create a list of intervals defining regions accessible in one or both cMNs. This final master list of intervals was used to narrow the total genomic search space for each disease group, with only variants contained in the regions specific to the cMN(s) of interest being retained.

### Modes of Inheritance

In order to leverage pedigree information, we first stratified our 270 pedigrees into 7 major disease categories that shared cell type specific aetiology (CFEOM, FNP, DRS, CFP, Moebius, Ptosis, Ptosis/MGJWS). We further stratified these pedigree groups into subgroups based on 4 inheritance/phenotype patterns (familial/syndromic; familial/isolated; trio/syndromic; trio/isolated). We incorporated inheritance by only retaining variants that matched appropriate mode(s) of inheritance in at least one family in a given subgroup. For example, for trios we searched variants obeying *de novo*, dominant (if either parent was affected), compound heterozygous, and/or homozygous recessive modes of inheritance. For *de novo* variants, we used Hail’s likelihood-based caller (https://github.com/ksamocha/de_novo_scripts). For familial cases, we manually inspected each pedigree structure and specified custom variant searches based on plausible modes of inheritance, including *de novo*, dominant, compound heterozygous, homozygous recessive, and dominant with incomplete penetrance. In the case of compound heterozygous variant configurations affecting non-coding elements, we defined each scATAC peak as our unit of heredity. Within this framework, one variant in a peak had to be inherited from an unaffected father, and a different variant in the same peak had to be inherited from an unaffected mother. Finally, we performed cohort-level filtering by eliminating any rare candidate variants that were also present in any unaffected individuals in the cohort (for dominant / *de novo* searches) or that were present in a homozygous state in any unaffected individual (for recessive searches). We removed one outlier pedigree which had an excessive number of candidate variant calls.

For SV genetic interpretation, we performed inheritance based searches for dominant/*de novo* modes of inheritance in the appropriate pedigrees, using the same custom search parameters as described for the SNV/indel framework. We identified all *de novo* and inherited variants overlapping disease-relevant peaks for each eligible pedigree using the *findOverlapPairs()* function from the GenomicRanges package.

For TE genetic interpretation, we imported the list of TEs called with xTEA^149^ into Hail as a MatrixTable. We performed inheritance-based searches for dominant/*de novo* modes of inheritance, again using the same custom search parameters as described for the SNV/indel framework. We converted the TE MatrixTable from hg19 coordinates to hg38, and filtered out calls with invalid/unknown contigs, and only included highest confidence calls (Feature info = “two_side_tprt_both”). We applied estimated gnomAD AF thresholds of 0.01 and 0 for dominant inherited and *de novo* alleles, respectively. We used the same cell type-specific peak interval/disease group combination described above but added +/-15bp padding to each peak to account for uncertainty in the insertion point.

To identify multi-hit peaks, we aggregated candidate variant results within each cell type/disease pairing by peak and selected for any peaks with SNVs/indels and/or SVs present in ≥ 2 families. For multi-hit tabulation, we excluded any SVs > 100 kb or with clear coding etiology. Variants within multi-hit peaks were required to obey the same broad mode of inheritance (i.e., dominant or recessive). In addition, dominant and recessive multi-hit variants could not be present in any unaffected individual across the cohort in the heterozygous and homozygous configuration, respectively. Candidate variants in any previously solved pedigrees were excluded from final tabulation^19,21,22,27,34,87,88,90,92,155–161^.

### Permutation testing

To assess the statistical significance of the results that lie within the regions drawn from scATAC sequencing of developing cranial motor neurons, we performed permutation tests to determine whether the regions corresponding to specific cranial motor neurons were enriched for variants. We analyzed dominant inherited and de novo variants separately.

First, we performed a search to find variants using the same thresholds for frequency, conservation, quality, and inheritance, but without limiting the search space to only genomic intervals defined in the scATAC peaks. We then split these results by disease group based on the phenotype of the family to create the genome-wide distribution of candidate variants for each disease group. After examining the distribution of the number of genome-wide de novo variants per individual after filtering for thresholds, we removed four individuals from the results due to existing significantly outside of the distribution (with the threshold drawn at >75 de novos per individual).

We then conducted permutation tests on each disease group, using regioneR.^162^ We used the original set of genomic locations from the cranial motor neuron(s) scATAC data to randomly generate a new list of peaks. The new list of randomly generated peaks was restricted to the same peak sizes and number of peaks as the original list, and could not overlap. We used the hg38 masked genome from BSGenomes in order to restrict the locations where the randomized peaks could be located. We then counted the number of variants within these new regions. This process was repeated for 5000 iterations for each disease group for both de novo and dominant inherited variants.

### ddPCR copy number validation

We performed ddPCR droplet generation and droplet reading using the QX200 droplet digital PCR system with Biorad ddPCR Supermix for Probes (Bio-Rad #186-3010). We performed copy number genotyping for non-coding element hs2757 in pedigrees S190 and S138 using ddPCR Copy Number Assay (Bio-Rad dHsaCNS845311073) and TaqMan Copy Number Reference Assay, human, TERT (Life Tech 4403315) as an internal control. We used the following thermocycler protocol: 1 x [95°C for 10 min]; 40 x [94°C for 30s, 60°C for 1 min]; 1 x [98°C for 10 min], 1 x [4°C hold]. Genotyping was performed in duplicate for all samples.

### Convolutional neural network training and prediction

We generated accessibility predictions using *Basenji*^110,162^ after training the network with mouse motor neuron scATAC-seq data. We generated separate predictions for each biological replicate (32 replicates total). To preprocess scATAC-seq data before training the neural network, we first generated bigwigs from the scATAC-seq bam files using mm10 as the reference FASTA. We clipped bigwig coverage at 150 to trim outliers. We generated training, validation, and test sequences with a split of 80% training sequences, 10% validation, and 10% test. We identified regions that should not be included in training sequences with a bed file containing regions that were hard masked in the mm10 fasta file combined with the Encode denylist. The mm10 FASTA file was filtered to only include chromosomes 1-19, X, and Y.

We trained the network retaining the model architecture from the original Basenji manuscript, with seven dilated layers. For this work, the dense output layer contained 32 units (one for each sample). Training was stopped when the correlation coefficient for validation predictions vs. validation experimental data failed to improve after 12 iterations (patience = 12), and the weights from the best iteration were saved as the final model. The complete architecture and list of hyperparameters can be found at https://github.com/arthurlee617/noncoding-mendel under *params.json*.

Using this trained network, we generated SNP activity difference (SAD) scores for each human candidate variant by calculating the total difference in predicted reference vs. alternate coverage over a 131,072 bp window centered about each variant site (hg38). Here we made the implicit assumption that a network trained on mouse accessibility data was portable across species within the same cell type^110,163^. We also included four solved CFP pathogenic variants as truth data. For ease of interpretation, we converted all SNV predictions from raw counts differences to Z-scores, which fit a normal distribution. To calculate Z-scores for individual candidate indels, we used the SNV derived scores for our null distribution.

### Non-coding CRISPR mice and binomial ATAC

We performed scATAC-seq for GFP-positive cMN7 e10.5 from two CRISPR-mutagenized mouse lines (*cRE2^Fam^*^4^*^/Fam^*^4^ and *cRE2^Fam^*^5^*^/Fam^*^5^) corresponding to human non-coding pathogenic variants described previously. *cRE2^Fam^*^5^*^/Fam^*^5^ is reported previously, corresponding to the pathogenic SNV (chr6:88224892A>G) mouse line^163^. *cRE2^Fam^*^4^*^/Fam^*^4^ (chr6:88224893C>T) was mutated on a C57Bl6 background via CRISPR-Cas9 homology directed repair at the Boston Children’s Hospital Gene Manipulation & Genome Editing Core and subsequently crossed onto the mixed *Isl^MN^*:GFP line described above. For each mutant line, we generated two biological replicates (4 replicates total) on embryos from [homozygous mutant x homozygous mutant] timed matings and compared to our wildtype cMN7 e10.5 replicates. For *ad hoc* comparison across these samples, we performed iterative LSI dimensionality reduction and batch correction using *Harmony*^164^ and normalised coverage by log_10_(nfrags). We note that *cRE2^Fam^*^4^*^/Fam^*^4^ also harbours an off-target C>T variant 54bp downstream from the target site (i.e., in addition to the on-target variant). This off-target nucleotide is not mutated in any affected samples. However, we do not explicitly exclude the possibility that this off-target variant contributes to the difference in *cRE2^Fam^*^4^*^/Fam^*^4^ accessibility relative to wildtype. For binomial ATAC, we performed [wildtype x homozygous mutant] timed matings for GFP-positive cMN7 from the e10.5 *cRE2^Fam^*^5^*^/Fam^*^5^ line, again across two biological replicates.

To test the *cis* effects of the mutant allele on accessibility, we tabulated reference versus mutant allele counts and performed a two-sided exact binomial test:

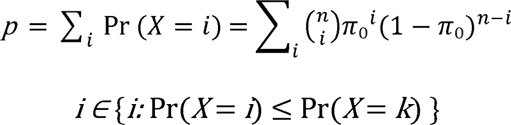

where the number of trials, *n* corresponds to sequencing coverage, the number of successes, *k* corresponds to reference allele count, and the expected probability of success, *π_0_* corresponds to the expected sampling probability of the reference allele under the null hypothesis *H*_0_: *π* = 0.5.

## Data availability

All data generated in this work are available through the Gene Expression Omnibus accession number **GSExxxxxx.**

## Code availability

Custom code to perform analyses from this work is available at https://github.com/arthurlee617/noncoding-mendel.

